# Randomised clinical trial of long term glutathione supplementation offers protection from oxidative damage, improves HbA1c in elderly type 2 diabetic patients

**DOI:** 10.1101/2021.04.27.21256157

**Authors:** Saurabh Kalamkar, Jhankar Acharya, Arjun Kolappurath Madathil, Vijay Gajjar, Uma Divate, Sucheta Karandikar-Iyer, Pranay Goel, Saroj Ghaskadbi

## Abstract

Sustained hyperglycemia in type 2 diabetes is associated with low levels of the endogenous antioxidant glutathione (GSH), which leads to oxidative stress and tissue damage. It is therefore important to investigate whether oral GSH supplementation would restore GSH levels, and also help improve redox state and glycaemia in diabetic individuals. We conducted a pragmatic clinical trial to assess effectiveness of oral GSH supplementation along with anti-diabetic treatment. 104 non-diabetic and 250 diabetic individuals were recruited. All the 250 diabetic individuals were on anti-diabetic therapy; of these 125 were given oral GSH supplementation additionally for a period of six months. Fasting and PP glucose and insulin, HbA1c, GSH and the oxidative DNA damage marker 8-OHdG were measured at the recruitment time and after three and six months of supplementation. GSH supplementation increased blood GSH and decreased 8-OHdG significantly within three months in diabetic individuals. It also reduced HbA1c within three months and stabilized it thereafter in diabetic population overall. Patients above 55 years benefited more as evidenced by a significant decrease in HbA1c and 8-OHdG and an increase in fasting insulin. Data suggest that GSH supplementation can be used as an adjunct therapy to anti-diabetic treatment to achieve better glycemic targets, especially in elderly population.

**Trial registered:** This study has been registered with Clinical Trials Registry-India (CTRI/2018/01/011257)

**Novelty bullets:**

- Oral GSH supplementation replenishes body stores of GSH and significantly reduces oxidative DNA damage in diabetic patients over six months.
- HbA1c is lowered at three months and is stabilized thereafter.
- A significant decrease in HbA1c and elevated fasting insulin was observed in people older than 55 years.

## Introduction

Hyperglycemia accelerates the production of reactive oxygen species (ROS) and leads to substantial oxidative stress (OS) in diabetic patients (Brownlee, 2001). Hyperglycemia-mediated ROS production leading to oxidative damage to biomolecules, is in fact, proposed to be a common unifying mechanism of diabetic complications. ROS activates four different pathways namely, advanced glycation end products (AGEs) (Shinomara et al., 1998), polyol (Lee and Chung 1998), hexosamine (Kolm-Litty et al., 1998), and protein kinase C (Derubertis and Craven, 1994), all of which are involved in the development of diabetic microvascular complications (Brownlee, 2005). Therefore, one important strategy for alleviating diabetic complications could potentially be to scavenge hyperglycemia-mediated ROS in diabetic patients. Studies in animal models of diabetes have shown that treatment with antioxidants such as, N-Acetyl-L-Cysteine (NAC) (Kaneto et al., 1999; Haber et al., 2003), lipoic acid (Moini et al., 2002), and glutathione (GSH) (Aw et al., 1991; Ueno et al., 2002) or precursors of GSH, such as glycine and cysteine (Jain et al., 2009; El-Hafidi et al., 2018), partially improves the blood glucose levels, functionality of β-cells, insulin sensitivity and reduces diabetic complications. However, human clinical trials using antioxidants in patients with cardiovascular diseases and metabolic syndrome have not been conclusive (Johansen et al., 2005).

Clinical trials of GSH supplementation as an endogenous antioxidant have recently received a great deal of attention. GSH, a water-soluble tri-peptide synthesized from glutamate, cysteine, and glycine, is an important endogenous antioxidant required for maintaining redox homeostasis of the cell. It is synthesized by glutathione synthetase and utilized by GSH peroxidase and glutaredoxin to detoxify free radicals. In the process, it is converted to its oxidized form, GSSG which is converted back to GSH by glutathione reductase (Njalson and Norgren, 2007). Low GSH levels are known to be associated with several pathological conditions, such as cancer, arthritis, cardiovascular and neurodegenerative diseases, and diabetes (Nuttall et al., 1998; Townsend et al., 2003). Several reports, including work from our lab, have confirmed that GSH levels are significantly low in diabetic patients (Martian-Gallan et al 2003; Song et al., 2007; Acharya et al., 2014; Picu et al., 2017). GSH insufficiency can be due to increased exposure to oxidants, drugs, excess nutrients or decreased rate of synthesis of GSH arising from nutritional deficiency of its precursor amino acids (De Luca et al., 2001). Genetic factors such as SNPs in the genes coding for enzymes required for GSH synthesis may also result in decreased GSH levels. It is plausible that GSH supplementation may help in arresting the development of complications in type 2 diabetes by improving the redox state and minimizing the oxidative damage to the biomolecules.

Oral administration of GSH or its precursor amino acids has been shown to enhance body stores of GSH, in plasma as well as tissues (Aw et al.,1991; Favilli et al., 1997; Kariya et al. 2007) and protected against different pathological conditions. Richie et al. (2015) demonstrated its effectiveness in restoring body stores of GSH in a dose and time dependent manner. Several different forms of GSH, such as sublingual (Schmitt et al., 2015), orobucccal (Buonocore et al., 2016; Bruggeman et al., 2019) and liposomal (Sinha et al., 2018) have recently been reported to be effective in restoring body stores of GSH. Supplementation with glycine, a precursor of GSH, has also been shown to increase the rate of GSH synthesis in elderly individuals (Sekhar et al., 2011a). However, results from human clinical trials of oral GSH supplementation have often been contrasting and debatable. Allen and Bradley, (2011) reported that oral GSH supplementation in healthy adults did not change GSH levels and other biomarkers of OS. Whereas, Richie et al., demonstrated that oral GSH supplementation leads to significant increase in body stores glutathione (Richie et al., 2015). Discrepancies in the outcomes of these studies could be attributed to differences in the dose and duration of GSH, and measurement of GSH in human plasma instead of erythrocyte fraction, where GSH is known to be turned over rapidly.

Most studies of GSH supplementation have focused on restoring body stores of GSH. However, it is likely that the relationships of GSH supplementation with pathology are complex. We have previously demonstrated that controlling hyperglycemia over a period of two months increases blood GSH levels and reduces oxidative damage significantly, regardless of the anti-diabetic treatment prescribed to them (Acharya et al., 2014). More than a dozen markers of OS were measured; of these, GSH impressively correlated with glucose recovery, that is, it responded rapidly, within 8 weeks, to changes in glycosylated haemoglobin (HbA1c) (Kulkarni et al., 2014). This suggests that altering hyperglycemia rapidly results in changes in GSH. It is equally interesting to ask whether changes in GSH feed back into glycemia? It is well understood, for instance, that β-cells are poorly protected against OS (Lenzen et al.,, 1996; Tiedge et al., 1997), hence an improvement in the redox state might, plausibly, relieve stress on β-cells, and in turn assist glycemic control. A few studies have examined the effects of GSH infusion on improving glycemic parameters in diabetic patients. Paolisso et al. (Piolisso et al., 1992a) reported increased GSH, and total body glucose disposal; further, this effect was more pronounced in elderly individuals with impaired glucose tolerance (Piolisso et al., 1992b). On the other hand, Sekhar et al. (Sekhar et al 2011b) showed that infusion with glycine, a precursor of GSH, increased the rate of GSH synthesis and reduced lipid peroxidation in diabetic individuals; however, it did not lead to changes in HbA1c.

We hypothesize that oral GSH supplementation can (i) increase body stores, (ii) improve the systemic redox state to alleviate OS and, further (iii) protect β-cells against OS (iv) improve insulin secretion, and (v) help anti-diabetic treatment in restoring normoglycemia.

Here we have conducted a pragmatic prospective clinical trial to assess whether oral GSH supplementation improves body stores of GSH in diabetic patients. We further asked if the effect of GSH supplementation in combination with anti-diabetic treatment is cumulative, that is, it has an additional benefit in improving glucose control and minimising oxidative damage in diabetic patients. We serially measured concentrations of GSH, 8-hydroxy-2-deoxy guanosine (8-OHdG), an oxidative damage marker, and glycemic parameters in diabetic patients receiving GSH supplementation in addition to anti-diabetic treatment, and compared them with serial measurements in those receiving anti-diabetic treatment alone. We find that GSH supplementation not only improved body stores of GSH, and decreased oxidative damage to DNA but also helped stabilize lowered HbA1c in diabetic patients. We noted that this effect of GSH supplementation is more pronounced in elderly diabetic patients (age more than 55 years).

## Materials and methods

### Study design

We conducted a pragmatic clinical trial which is designed as a case-control cohort study to assess the effect of oral GSH supplementation on blood GSH levels and glucose homeostasis in diabetic patients.

### Study approval

This study was approved by the Institutional Ethical Committee of Jehangir Hospital Development Center, Pune (Dr. Ravindra Ghooi, Chairman, Ethics committee Jehangir clinical development center ECN-ECR/352/Inst/NIH/2013); Institutional Biosafety Committee of SPPU (Bot/27A/15), Pune; and Institutional Ethical Committee of IISER, Pune. Signed informed consent was obtained from all the subjects at the time of enrollment in the study after explaining the purpose and nature of the study. All participants in this study were de-identified using a numbered code. This study is registered with Clinical Trials Registry-India (CTRI/2018/01/011257).

### Inclusion/Exclusion criteria for study participants

We recruited healthy non-diabetic controls (n=104) (Control group) with HbA1c < 6.5%, and known type 2 diabetic subjects (n=250) with HbA1c ≥ 6.5% (ADA, 2016) of either sex between age of 30-70 years visiting Jehangir Hospital and Iyer clinic, Pune. Pregnant women, individuals with excessive alcohol intake, heavy smokers, individuals with any clinical infection or with a history of a recent cardiovascular event, and those receiving antioxidants were excluded from the study. Body weight, height, anti-diabetic treatment and family history of diabetes were noted for each subject.

### Recruitment and randomisation for GSH intervention

We recruited known diabetic subjects (n=250) who were already on anti-diabetic regimen and randomly categorized them into two groups: 125 diabetic patients who were advised to continue with their anti-diabetic regimen as prescribed by physician (Group D), and the other 125 diabetic patients who were given 500 mg glutathione (Jarrow Formulas, USA) supplementation once daily in addition to their anti-diabetic treatment (Supplemental Table S1) for a period of six months (Group DG) (Fig. 1). At the time of randomisation, concentrations of covariates fasting and PP glucose and insulin, HbA1c, GSH and oxidized glutathione (GSSG), and 8-OhdG were not available, and therefore, did not influence the assignment of diabetic patients in D or DG groups. Patients in the DG group were issued two bottles of GSH (each bottle containing 60 capsules of 500 mg each) and were called back after three months to collect an additional bottle for the remaining duration. During the tenure of the study compliance to medical treatment by patients of D and DG group and consumption of GSH by patients of the DG was emphasized by maintaining continuous communication between the physician and patients. Out of 125 diabetic patients in D and DG group, 23 were lost to follow-up in the D group and 21 in DG group for not complying with the treatment regimen. We also recruited healthy non-diabetic control subjects who were followed for six months during which they were advised to continue with their regular diet and exercise regimen. Blood samples were collected at the time of enrollment 0 (alpha (α) visit), 3 (beta (β) visit) and 6 (gamma (γ) visit) months after the date of enrollment.

**Fig. 1.**
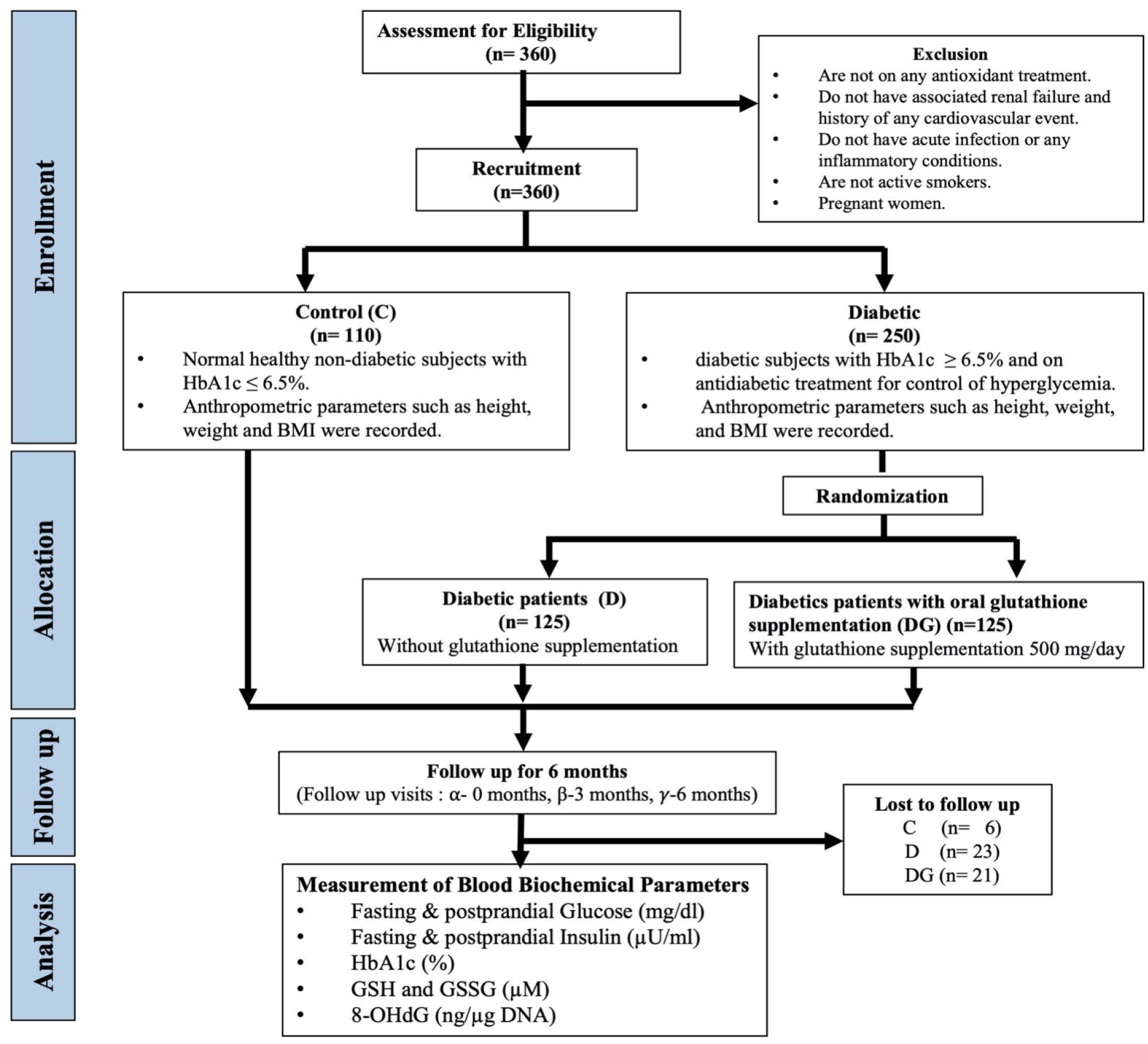
Flowchart for study design.

### Sample collection

At each visit, a total of 10 ml fasting and postprandial (PP) blood samples were collected from all the subjects by Golwilkar Metropolis, Pune-411004, in 5 and 2 ml vacutainers containing EDTA for collecting plasma and whole blood samples, and 3 ml vacutainers without EDTA for collecting serum. Whole blood samples were centrifuged at 4000 rpm for 10 mins to separate plasma and erythrocyte fraction. Plasma was separated and stored at −80°C (Thermo Fisher Scientific, USA) for further analysis.

### Outcomes

Primary outcome measures include GSH and HbA1c. Secondary outcomes include GSSG, fasting glucose (FPG) and insulin (FPI), PP glucose (PPG), and insulin (PPI), and 8-OHdG.

### Estimation of blood biochemical parameters

Measurement of FPG, PPG, FPI, PPI, and HbA1c was performed on an automated analyzer at Golwilkar Metropolis, Pune, by following CLSI (Clinical and Laboratory Standards Institute, USA) guidelines. Erythrocyte fraction was then used for preparation of hemolysates by washing it twice with cold saline (9.0 g/L NaCl) and hemolyzed by adding ice-cold ultrapure water (MilliQ plus reagent grade; Millipore, Bedford, MA, USA) to get 50% diluted hemolysates (Acharya et al., 2014) which were stored at −80°C for further analysis.

### Estimation of GSH and GSSG

Reduced and total glutathione content in erythrocyte hemolysate was estimated using Glutathione assay kit (Cayman Chemical, USA). This kit follows DTNB (5,5’-dithio-bis-2 nitrobenzoic acid, Ellman’s reagent) method for estimation of GSH (Baker et al., 1990) where DTNB reacts with reduced GSH yielding yellow-colored 2-nitro-5-thiobenzoate, which is read at 405 nm, on ELISA reader.

Briefly, 50 µl of erythrocyte lysate was deproteinized using an equal volume of metaphosphoric acid at 4°C. After vigorous vortexing, the resulting mixture was centrifuged at 2000 g for 2 mins at 4°C. The supernatant was separated and aliquoted in two parts and used for estimation of total GSH (TGSH) and GSSG. pH of the samples was adjusted to 8 by the addition of triethanolamine (5 µl/100 µl of the sample). One of the aliquots was diluted 50 times with 1x MES buffer (0.4 M (N-morpholino) ethanesulphonic acid, 0.1 M phosphate buffer and 2 mM EDTA) and used for estimation of TGSH. In the second aliquot, 2 µl of vinyl pyridine was added and diluted 25 times with 1x MES buffer and 50 µl of this sample was used for estimation of GSSG. Both the aliquots were then incubated for 1 hour at room temperature. The reaction was started by adding 150 µl assay cocktail containing 11.25 ml MES buffer, 0.45 ml cofactor mixture containing NADP^+^ and glucose-6-phosphate, 2.1 ml enzyme mixture containing glutathione reductase and glucose-6-phosphate dehydrogenase, 2.3 ml water and 0.45 ml DTNB. Increase in TNB formation was determined by measuring absorbance at 405 nm at 5 min interval for 30 mins. GSSG was used as a standard for estimating the concentration of TGSH and GSSG in samples. Absorbance values of samples and standard curve (0, 0.5, 1.0, 2.0, 4.0, 8.0, 12.0, and 16.0 µM) were plotted as a function of time and slope for each sample was calculated. This was called i-slope. i slope of each concentration of standard was plotted against the concentration of GSH and the slope of this curve was called as f-slope. Values for total GSH and GSSG were calculated by using the formula given below. GSH concentration was determined by subtracting GSSG from total GSH.

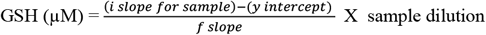

### Estimation of 8-OHdG

DNA was isolated from whole blood by standard phenol-chloroform-isoamyl alcohol extraction method, quantitated on nanodrop and concentration of 8-OHdG was determined by competitive enzyme-linked immunosorbent assay following the protocol of Modak et al. (2009) and expressed as nanogram 8-OHdG/µg DNA.

### Statistical methods

Biochemical parameters of subjects in Control, D and DG groups at the first visit were represented using the descriptive statistics (Median, 25th percentile, 75th percentile). All intra-group and inter-group comparisons of biochemical parameters at different visits were performed using permutation tests, using the “Coin” package in R (Hothorn et a;., 2006). Statistical significance was set at p-value < 0.05. The results of permutation tests were confirmed with two-sample, two-sided t-tests, and are reported in Supplemental sections. Effect size analysis was used to quantify the difference between 6-month biochemical changes from α to γ visit in D and DG groups. Statistical analysis and effect size calculations were repeated between subgroups in D and DG consisting of elderly subjects with age more than 55 years. All effect size calculations and parametric t-tests for comparisons were carried out using Matlab version 2019.

### Effect size between change in the biochemical levels of D, and DG group individuals

Biochemical measurements at α, β and γ visits of the D group subjects are represented by variables 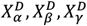 and measurements from DG group subjects by 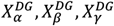, respectively. The biochemical variables, X are HbA1c, FPG, FPI, PPG, PPI, GSH, GSSG, and 8-OHdG. Changes from α to γ visit (6-months) in D were estimated by taking their paired differences as 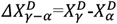, with a group-wise mean of paired difference, 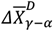. Similarly, 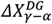 represents the paired changes from α to γ visit (6 months) with a group-wise mean, 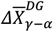 in DG.

The effect size of 6-month changes in the concentration of a particular biochemical variable, X, between D and DG groups are estimated using **Cohen’s d** (Cohen, 1988) as

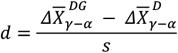

where pooled standard deviation of biochemical changes in D and DG, *s*, is given by

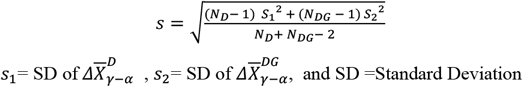

Here *N*_*C*_, *N*_*D*_ and *N*_*DG*_ are the number of individuals in Control, D and DG groups respectively. Cohen (1969) described an effect size of 0.2, 0.5, and 0.8 as “Small”, “Medium”, and “Large” effect respectively, and Sawilowsky (2003), classified an effect of size 1.2 as “Very large” and 2 as a “Huge” effect.

## Results

### Baseline characteristics of study groups

The study population included diabetic subjects with a mean age of 54 years and BMI 26.9 kg/m^2^ and the control group included individuals with a mean age of 41 years and BMI 26 kg/m^2^. D group comprised of 57 males and 45 females, DG group contained 49 males and 55 females, and the control group comprised of 62 males and 42 females. Baseline characteristics of subjects in each group are presented in Table 1.

**Table 1.**
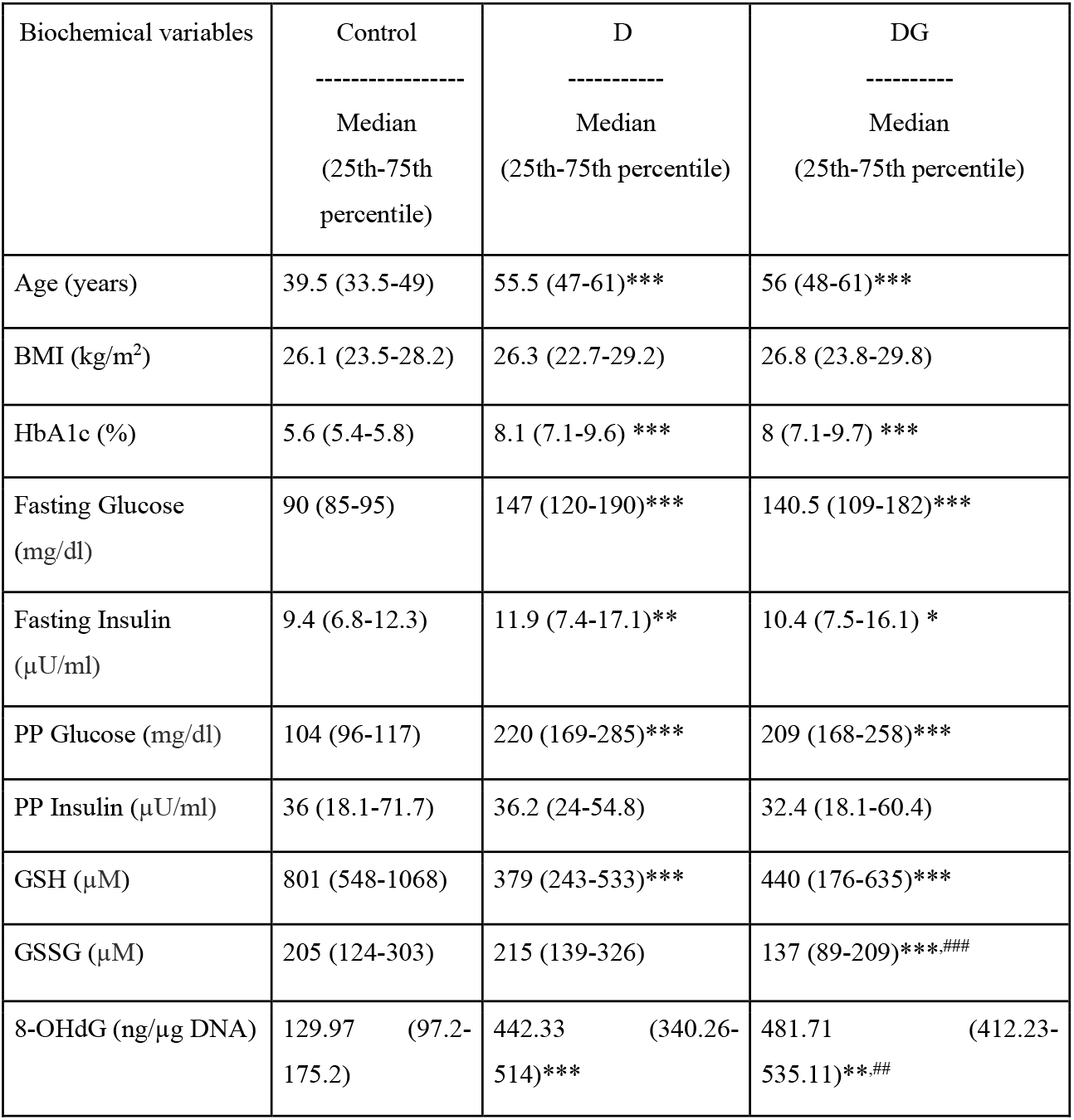
Baseline characteristics of Control, D, and DG groups. Data from each group at the α visit are presented here as median and interquartile ranges (25th percentile-75th percentile). * indicates the significance of the comparison between baseline measurements of Control versus D or Control versus DG groups. Significance levels are *p < 0.05, **p < 0.01, ***p < 0.001. (See Table S3 for alternate comparisons using two-sample, two-sided t-tests). # indicates the significance of the comparison between D and DG using two-sample permutation tests (p <0.05).

Concentrations of FPG, PPG, FPI, HbA1c, and 8-OHdG were significantly high and that of GSH was significantly low in D and DG compared to Control (p<0.001, all parameters). Levels of PPI in D and DG were not found to be significantly different compared to the control group (Table 1).

We did not observe any significant differences in the levels of FPG, PPG, FPI, PPI, HbA1c, and GSH within D and DG groups, thus, confirming covariate balance in the two groups at baseline. The concentration of GSSG, and 8-OHdG were found to be significantly different (p<0.05) between D and DG (Table 1).

### Oral GSH supplementation increases erythrocyte GSH, GSSG and decreases oxidative damage to DNA but does not alter glycemia in diabetic patients over a period of six months

Our study estimates the effect of oral GSH supplementation on erythrocyte GSH concentrations over a period of six months in diabetic patients. GSH levels increased significantly over a period of six months, from α to γ visit in both DG (p<0.001) and D (p<0.001) groups, while they remained unchanged in the Control. We further estimated the effect size of GSH supplementation within the diabetic groups: A “Large” effect (Cohen’s d = 1.01; p<0.001) indicated that the increase in GSH is significantly high in DG compared to D (Fig. 2). GSSG was similarly increased in DG compared to D (Cohen’s d=0.61, p<0.001).

**Fig. 2.**
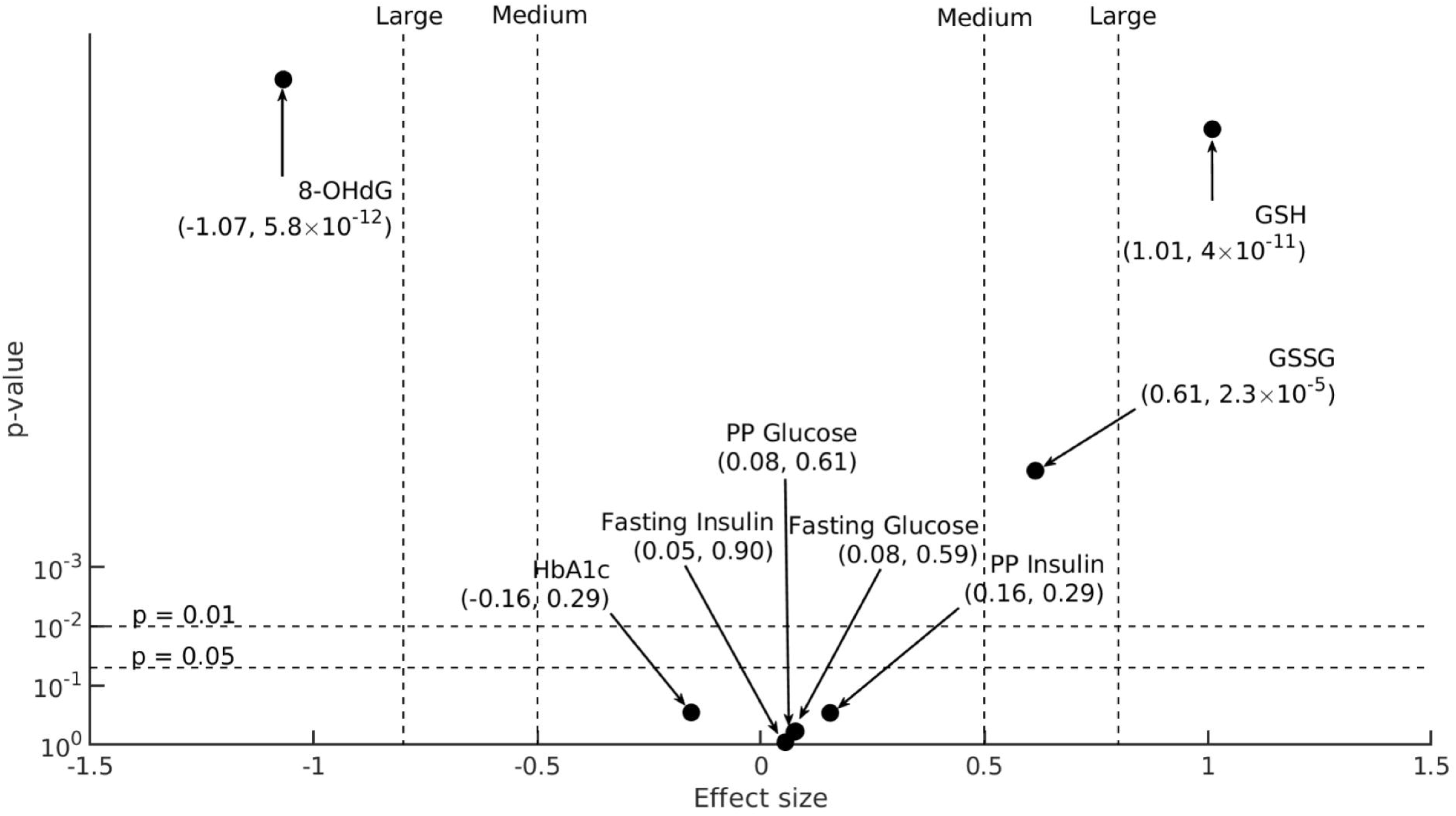
The effect size of changes in blood biochemical parameters. 6-month changes in biochemical parameters (X= HbA1c, FPG, FPI, PPG, PPI, GSH, GSSG, and 8-OHdG) D and DG groups, 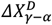 and 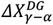, respectively (sample sizes for HbA1c, FPG, FPI are: *N*_*D*_ = 90, *N*_*DG*_ = 104, for PPG and PPI:*N*_*D*_= 90, *N*_*DG*_ = 102, for GSH and GSSG:*N*_*D*_= 89, *N*_*DG*_ = 101, for 8-OHdG: *N*_*D*_ = 91, *N*_*DG*_ = 104), are compared here on a statistical significance (y-axis) versus effect size (x-axis) plot. Effect size (Cohen’s d) calculated between 6-month changes in the concentration of biochemical variables, X, in D and DG (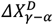 and 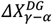) are denoted on the x-axis. The p-value of comparison between the group-wise means of 6-month changes in X’s concentration, 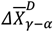 and 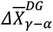 (Supplemental Table S4), obtained using two-sample permutation tests are denoted on the y-axis (Supplemental Figure S1 for an alternative version of this figure using two-sample, two-sided student’s t-tests instead). Two horizontal dotted lines denote 95% and 99% significance levels. Effect size takes either a positive or negative sign based on the direction of change: a positive effect size increases towards the right and a negative effect towards the left. Vertical dotted lines represent different classifications of effect size. In particular, “Medium” effects are labeled at 0.5 and −0.5, and “Large” effects at 0.8 and −0.8.

We also observed a significant decrease in the concentrations of 8-OHdG from α to γ visit in DG (p<0.001) but not in D and Control groups (p>0.05). GSH supplementation resulted in a “Large” effect (Cohen’s d = −1.07; p<0.001) indicating that 8-OHdG decreased significantly in DG compared to D (Fig. 2).

We then analyzed the effect of oral GSH supplementation on glycemic parameters, namely HbA1c, FPG, PPG, FPI, and PPI in diabetic patients. We observed that HbA1c levels decreased significantly over six months in both D and DG, however, the extent to which HbA1c levels decreased in DG was comparable to D group as indicated by a small Cohen’s d = −0.16 (p>0.05) (Fig. 2). FPG, PPG, FPI, and PPI decreased over a period of six months in D and DG, however, changes in DG were comparable to those in D (p>0.05, Cohen’s d<0.2, all parameters). Mean values for biochemical variables at 0, 3 and 6 months for the study groups are detailed in Supplemental Table S2.

Overall, our results indicate that GSH supplementation leads to a significant increase in the erythrocyte GSH and GSSG and decrease in the 8-OHdG concentrations in diabetic patients. However, glycemic parameters change in D and DG to similar extents.

Next, we investigated whether the effect of oral GSH supplementation on the above mentioned biochemical parameters is accomplished rapidly, say, within the first three months of GSH supplementation, and stabilized thereafter, or whether their levels change gradually over a period of six months. We, therefore, examined changes in the concentration of FPG, PPG, FPI, PPI, HbA1c, GSH, GSSG, and 8-OHdG serially from 0 to 3 and 6 months in each of the three groups. These results are described in the following section.

### Oral GSH supplementation enhances erythrocyte GSH concentration in diabetic subjects within three months

Fig. 3 a and b show serial changes in the concentrations of GSH, and GSSG, respectively, from α to β and γ visits in the three study groups. In Control, the concentrations of GSH and GSSG remained unchanged over a period of six months. GSH supplementation in DG led to a significant increase in GSH within the first three months (p<0.001) and remained stable thereafter up to six months (Fig. 3a). In the D group, on the other hand, GSH increased gradually from 0 to 3 and 6 months. In the DG group (Fig. 3b), GSSG concentration also increased significantly within the first three months (p<0.001), and did not change further. In D, on the other hand, GSSG remained unchanged during the study period (p>0.05).

**Fig. 3.**
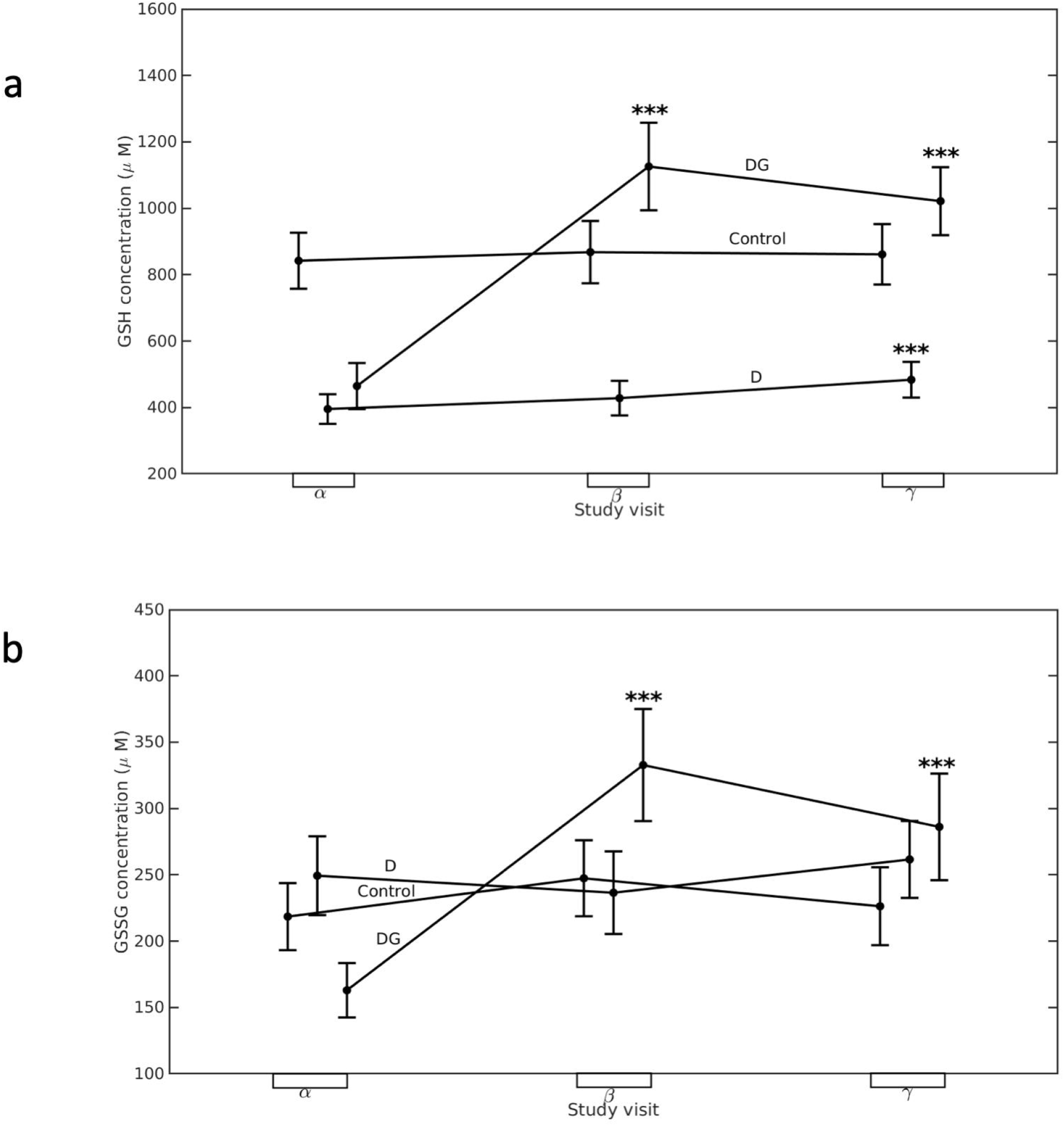
Longitudinal changes in the concentration of (a) GSH and (b) GSSG in different groups. Mean (black dots) and 95% confidence interval (whiskers) of (a) GSH and (b) GSSG concentrations at α visit (*N*_*C*_ = 104, *N*_*D*_ = 100, *N*_*DG*_ = 101), β visit (*N*_*C*_ = 104, *N*_*D*_ = 100, *N*_*DG*_ = 101), and γ visit (*N*_*C*_ = 104, *N*_*D*_ = 89, *N*_*DG*_ = 101) are shown here for Control, D and DG groups. Significance levels (*) displayed above β, and γ visits denote the comparisons with *α* visit using permutation tests. Significance levels are *p < 0.05, **p < 0.01, ***p < 0.001 for respective comparisons. (See Supplemental Fig. S2 for alternative comparisons using paired sample t-tests instead and Supplemental Table S6 for p-values of comparisons).

Thus, oral GSH supplementation in diabetic patients increased erythrocyte GSH concentration significantly within three months and stabilized it thereafter. On the other hand, in D, anti-diabetic therapy alone led to a small increase in GSH but not in GSSG.

### Oral GSH supplementation reduces 8-OHdG concentrations in diabetic subjects

Further, we evaluated the effect of GSH supplementation on serial changes in 8-OHdG in diabetic patients. In Control, its concentrations remained unchanged over a period of six months. GSH supplementation in diabetic patients led to a significant decrease in 8-OHdG within the first three months and it continued to reduce significantly thereafter (p<0.001) (Fig 4a). However, in the D group, its concentrations did not change significantly throughout the study period.

**Fig. 4.**
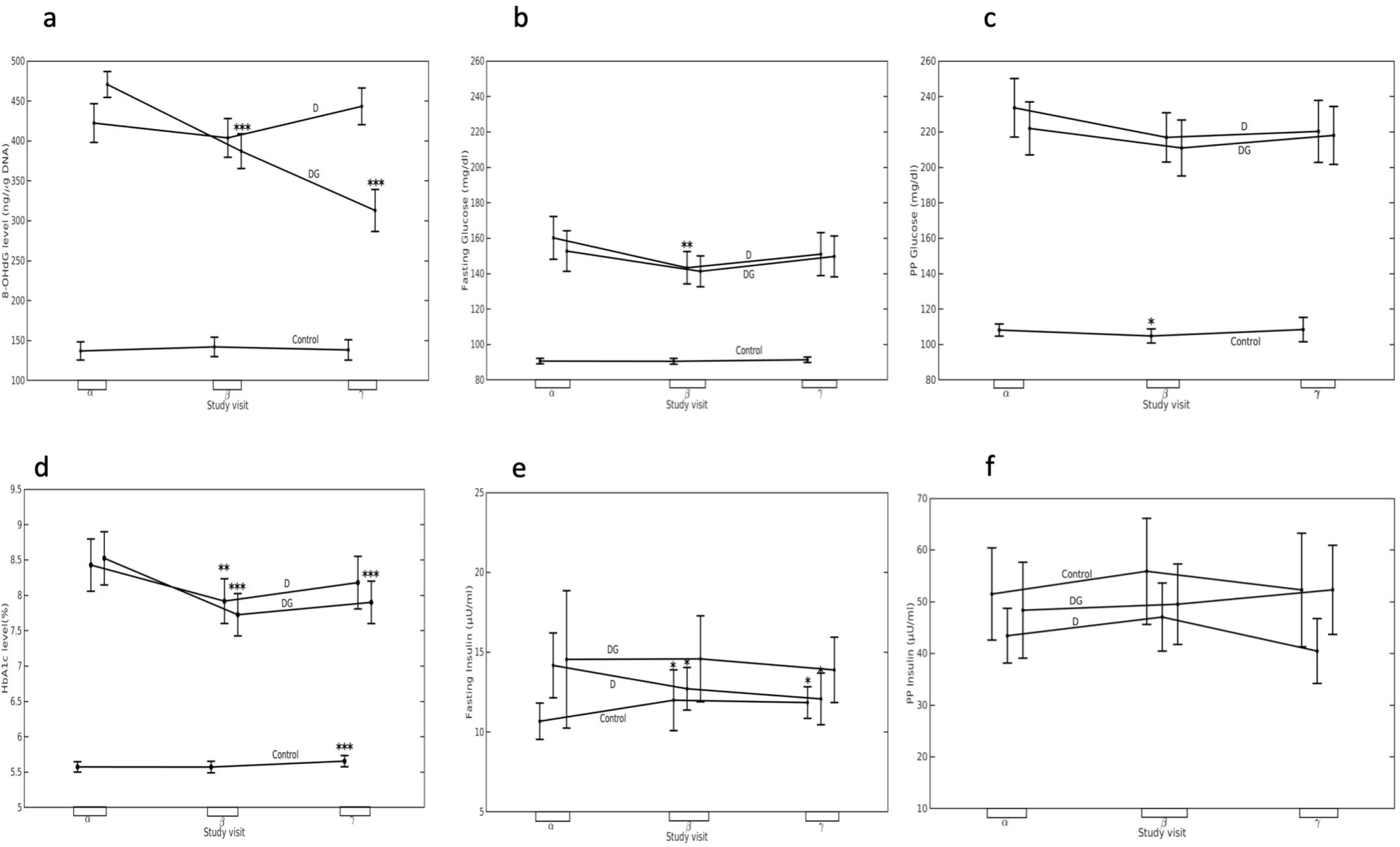
Longitudinal changes in glycemic parameters. Mean (black dots) and 95% confidence interval (whiskers) of (a) 8-OHdG, (b) FPG, (c) PPG, (d) HbA1c, (e) FPI, and (f) PPI concentrations at α (sample sizes for 8-OHdG, HbA1c, FPG, FPI, PPG are: *N*_*C*_ = 104, *N*_*D*_=102, *N*_*DG*_ =104; for PPI:*N*_*C*_ = 103, *N*_*D*_=102, *N*_*DG*_ =104; and for GSH, GSSG:*N*_*C*_ = 104, *N*_*D*_ =100, *N*_*DG*_ =101), β (for HbA1c, FPG and FPI:*N*_*C*_ = 104, *N*_*D*_=102, *N*_*DG*_ =103; for PPG:*N*_*C*_ = 103, *N*_*D*_=102, *N*_*DG*_ =103; for PPI:*N*_*C*_ = 103, *N*_*D*_=101, *N*_*DG*_ =102, for 8-OHdG: *N*_*C*_ =104, *N*_*D*_=102, *N*_*DG*_ = 104 and for GSH, GSSG:*N*_*C*_ = 104, *N*_*D*_ =100, *N*_*DG*_ =101), and γ visit (for HbA1c, FPG, FPI:*N*_*C*_ = 104, *N*_*D*_=90, *N*_*DG*_ =104; for PPG and PPI:*N*_*C*_ = 104, *N*_*D*_=90, *N*_*DG*_ =102, for 8-OHdG: *N*_*C*_ =104, *N*_*D*_=91, *N*_*DG*_ =104 and for GSH, GSSG:*N*_*C*_ = 104, *N*_*D*_ =89, *N*_*DG*_ =101 are shown here for Control, D, and DG groups. Significance levels (*) displayed above β and γ visits denote the comparisons with *α* visit using permutation tests. Significance levels are *p < 0.05, **p < 0.01, ***p < 0.001 for respective comparisons. (See Supplemental Fig. S3 for alternative comparisons using paired sample t-tests instead).

### Serial changes in HbA1c levels are stabilized by oral GSH supplementation in diabetic patients

We examined serial changes in the levels of FPG, PPG, HbA1c, FPI, and PPI in D and DG groups in greater detail. FPG levels lowered significantly within three months in D (p<0.01) and DG (p=0.05); however, it recovered to the baseline levels by the end of six months (Fig. 4b). PPG levels, on the other hand, did not change significantly in D and DG over a period of six months (p>0.05, all) (Fig. 4c). Thus, based on these results and Fig. 2, we conclude that GSH supplementation does not have any effect on FPG, and PPG levels in diabetic patients overall.

HbA1c levels rapidly decreased from 0 to 3 months in both D (p<0.01) and DG (p<0.001) (Fig 4d). Thereafter, HbA1c levels were maintained until 6 months in DG while they appear to have returned to the baseline level in D group.

FPI levels changed significantly from 0 to 3 and 6 months in D (p<0.05), while it remained unchanged in DG (p>0.05) (Fig 4e). PPI levels remained unaltered in D and DG throughout the study period (Fig. 4f).

Thus, the glycemic parameters, namely, FPG, PPG, FPI, and PPI do not change significantly in response to GSH supplementation. Taken together, oral GSH supplementation in diabetic patients appears to have “stabilizing effect” on HbA1c i.e., it decreases rapidly within three months and continues thereafter.

### Oral GSH supplementation significantly reduces HbA1c in a sub-group of diabetic patients, age>55 years

Earlier reports suggest that concentration of GSH decreases with age in healthy adults (Matsubara et al., 1991; Lang et al., 1992; Erdennal et al., 2002; Sekhar et al., 2011). Diabetes has also been reported to cause long term cardiovascular complications and congestive failure, which become marked with age. For instance, the Framingham study (Kannel, 1979) reported increased severity of chronic heart failures and related deaths in an age group above 55 years. Similarly, diabetes-related vascular complications in veterans (mean age = 60 ± 9 years) were also pointed out by Duckworth et al. (2009). Therefore, we believe it is important to assess the effect of GSH supplementation in elderly diabetic patients.

Diabetic patients in our study ranged from 31 to 78 years of age. The median age in these cohorts is about 55 years. This motivated us to use this age threshold to isolate the older sub-group. We then re-examined the effect of GSH supplementation in this sub-group diabetic population to assess whether they respond differently to oral GSH supplementation compared to the younger population.

Mean values for biochemical parameters GSH, GSSG, 8-OHdG, FPG, PPG, HbA1c, FPI, and PPI; and serial changes from 0 to 3 and 6 months in their concentrations in D (n=44) and DG (n=54) groups are shown in Supplemental Table S5 and Supplemental Fig. S5 and S6, respectively. Similar to results obtained for diabetic patients overall, the concentration of GSH and GSSG increased significantly over a period of 6 months in both D, and DG sub-groups (Supplemental Fig. S5). Changes in the mean GSH and GSSG levels over a period of 6 months in DG group (Fig. 5) was significantly higher compared to the D group (Cohen’s d = 1.14 and 0.67 for GSH and GSSG, respectively, p<0.001).

**Fig. 5.**
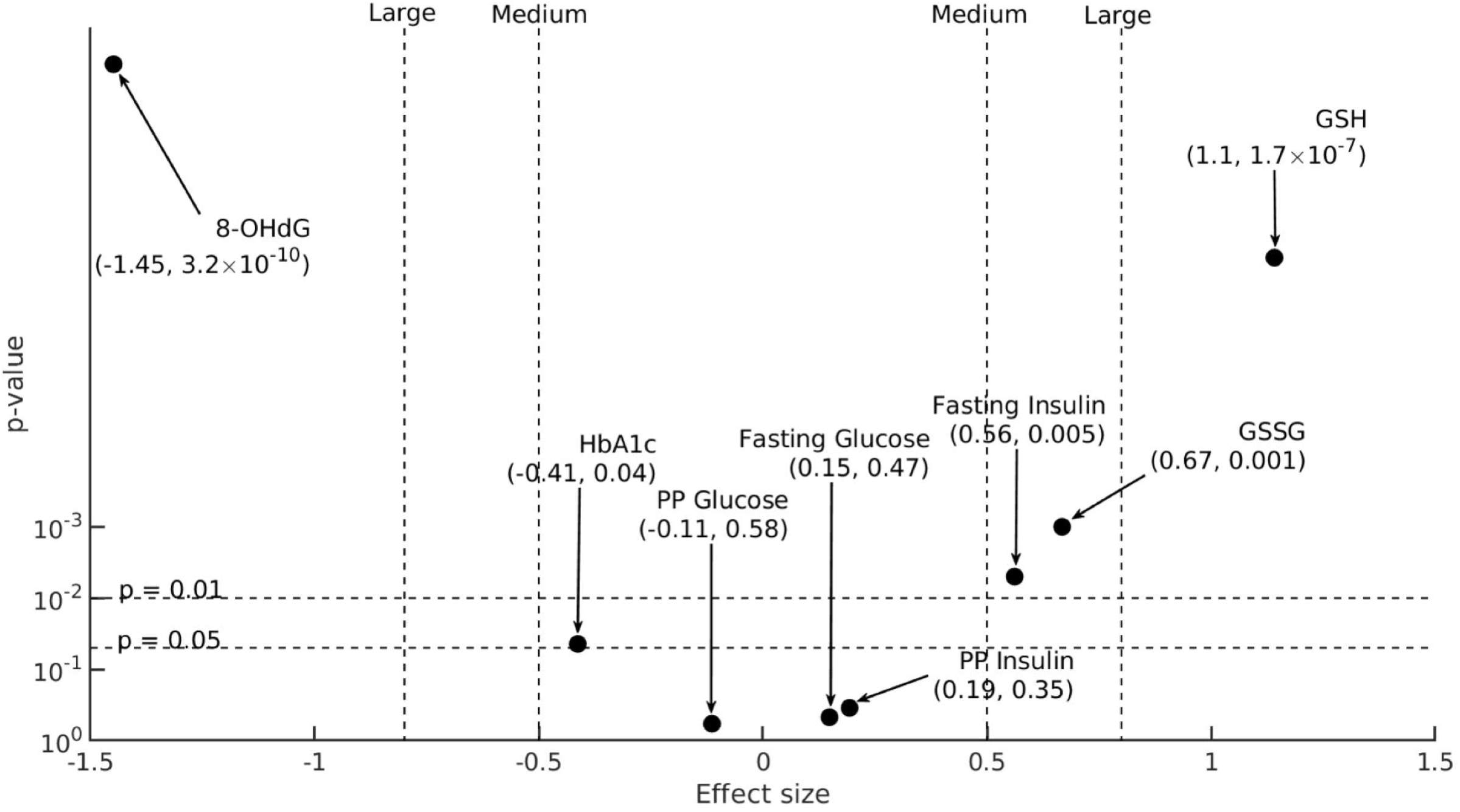
The effect size of changes in blood biochemical parameters of elderly diabetic patients. 6-month changes in biochemical parameters in D and DG sub-groups of elderly diabetic subjects (age > 55 years), 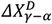 and 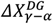, respectively (sample sizes for HbA1c, FPG, FPI are:*N*_*D*_ = 44, *N*_*DG*_ = 54, and for PPG, PPI, GSH, GSSG are:*N*_*D*_= 44, *N*_*DG*_ = 53, for 8-OHdG: *N*_*D*_= 45, *N*_*DG*_ = 54), are compared here on a statistical significance (y-axis) versus effect size (x-axis) plot. Effect size calculated between 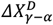 and 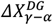, are denoted on the x-axis. The p-values of comparison between the subgroup-wise means of 6-month changes, 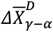 and 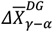 (Supplemental Table S8), obtained using permutation-tests are denoted on the y-axis (Supplemental Fig. S4 for alternative comparisons using two-sample, two-sided t-tests instead, and Supplemental Table S8 for p-values of comparisons).

Further, we also analysed the effect size of 8-OHdG concentrations in this elderly sub-group of diabetic patients on GSH supplementation. GSH supplementation resulted in a “Very large” effect (Cohen’s d = −1.45, p<0.001) indicating that 8-OHdG decreased significantly in DG compared to the D group over the period of 6 months (Fig. 5). This suggests that oral GSH supplementation in elderly diabetic population results in a significant reduction in the accumulation of oxidative DNA damage.

Next, we examined the effect size of blood glycemic parameters in response to oral GSH supplementation in the elderly sub-group of diabetic patients (Fig. 5). In contrast to the results observed in diabetic population overall, GSH supplementation in DG sub-group led to a significant reduction in HbA1c levels over a period of 6 months compared to D (Cohen’s d = - 0.41, p<0.05).

Interestingly, FPI levels also increased significantly in DG sub-group from α to γ visit compared to D (Cohen’s d = 0.56, p<0.05). GSH supplementation had a small effect on levels of FPG, PPG, and PPI in DG sub-group (Cohen’s d <0.2, p<0.05, all parameters). Mean changes in all biochemical parameters from α to γ visit in elderly diabetic patients of D and DG sub-groups are provided in Supplemental Table S7.

Thus, GSH supplementation in DG sub-group of elderly diabetic patients over a period of 6 months led to a significant increase in the erythrocyte GSH, GSSG, and FPI concentrations and reduced HbA1c and 8-OHdG levels significantly. This suggests that the elderly diabetic population responds to GSH supplementation in conjunction with anti-diabetic therapy better in terms of a significant reduction in HbA1c levels and accumulation of oxidative DNA damage compared to the diabetic population overall.

## Discussion

Sustained hyperglycemia is linked to an increase in OS which further results into risk of developing diabetic micro- and macrovascular complications (Brownlee, 2005). Interventions aiming at controlling hyperglycemia are the primary line of treatment for diabetic patients. It is interesting to ask if improving redox status by GSH supplementation can help counteract the deleterious effects of hyperglycemia-induced OS. GSH levels are known to be significantly low in diabetic patients, however, the bioavailability of oral GSH supplementation in humans is controversial (Allen and Bradley, 2011; Richie et al., 2015). Our study provides evidence that oral GSH supplementation not only improves body stores of GSH but significantly decreases accumulation of oxidative DNA damage and also helps increase the efficiency of regular anti-diabetic treatment in maintaining normoglycemia in diabetic patients.

GSH is known to be either transported in its intact form from the intestinal epithelial cells into blood lumen (Hagen et al., 1987; Hagen et al., 1990; Kovacs-Nolan et al., 2014) or broken down by gamma-glutamyltransferase (GGT) to its constituent amino acids (Hanigan, 2014). It is unclear whether GSH was either directly absorbed or was broken down into its constituent amino acids, and re-synthesized by glutathione synthetase. Additionally, we find a significant increase in the concentration of erythrocytic GSSG. This is possibly due to the conversion of erythrocytic GSH into GSSG, in line with previous reports, for instance Kovacs-Nolan et al. (2014) show that ^13^C-GSH administered to mice is rapidly converted to GSSG and accumulated in red blood cells and liver. Thus, oral GSH supplementation not only increases body stores of GSH but a fraction is stored as GSSG, and may be utilized for detoxification of free radicals. These results strongly suggest that GSH supplementation results in a systemic improvement of the redox state in diabetic individuals. Augmentation of antioxidant reserves, in the form of elevated GSH levels, also resulted in significant reduction in accumulation of oxidative DNA damage which is implicated in pathophysiology of diabetic complications.

HbA1c levels typically fluctuate despite regular anti-diabetic treatment in diabetic patients. We found GSH supplementation helped stabilize lowered HbA1c within three months. This effect was more pronounced in elderly patients, over 55 years of age. Other characteristics of the glycemic state, FPG, PPG, FPI, and PPI did not change in the full population of diabetic patients; however, interestingly we observed an increase in FPI levels in elderly diabetic patients. These results highlight that GSH supplementation distinguishes between the fine structure of glycemic changes, that is, while it influences long-term glycemia, as reflected in the stability of lowered HbA1c, short-term glycemic measurements, such as FPG and PPG, are unaltered. The exact mechanism by which GSH helps in maintaining normoglycemia in diabetic patients requires further investigation.

Preserving β-cell function is essential to glucose control in diabetic patients (DeFronzo and Abdul Ghani, 2011; Page and Reisman, 2013). It is crucial to maintain a healthy redox state of pancreatic β-cells as their ability to secrete insulin in response to glucose is dependent on intracellular and membrane thiols (Anjaneyulu et al., 1882). It is well established that β-cells are more vulnerable than other cells to hyperglycemia-mediated ROS due to their low antioxidant capacity and a poor ability to rectify oxidatively damaged DNA (Tiedge et al.,1997, Modak et al.,2009; Modak et al., 2014). Thus, one potential strategy towards improving glucose-stimulated insulin secretion is to provide antioxidant support to pancreatic β-cells. Enhancing extracellular GSH levels have been shown to improve β-cell response to glucose in rats (Ammon et al., 1989). These results were later confirmed by Paolisso et al.(1992a) who demonstrated that infusion with GSH enhanced β-cell response to glucose and consequently improved glucose tolerance in patients with impaired glucose tolerance. However, in routine clinical settings GSH infusion is not practical. We, therefore, sought to examine the effectiveness of oral GSH supplementation in ameliorating hyperglycemia in diabetic patients. Our results indicate that oral GSH supplementation supports anti-diabetic treatment in reducing hyperglycemia.

It is known that concentration of GSH declines with ageing (Matsubara and Machado, 1991; Sekhar et al 2011a), and this could be further aggravated in elderly diabetic patients. We indeed observed that elderly diabetic patients benefited more from GSH supplementation both in terms of reducing oxidative DNA damage and improving glycemic status.

Interestingly, we also observed a significant increase in the levels of FPI in these elderly patients. Similar observations have recently been reported by Zhang et al. (2019) in diabetic rats where ROS-induced damages to β-cell were prevented by administration of oral GSH. Islets isolated from type 2 diabetic cadaveric organ donors showed impairment in insulin secretion in response to glucose and increased level of oxidative damage markers; treating these islets with GSH led to an improvement in their functionality and also alleviated oxidative damage markers, suggesting that reducing OS in islets could be a potential target for treating type 2 diabetes (Del Guerra et al., 2005). We speculate that a systemic increase in GSH levels in diabetic patients resulted in significant reduction in oxidative DNA damage leading to an improvement in the insulin secreting ability of pancreatic β-cells and a concomitant reduction in HbA1c. However, these results need to be further validated in large clinical settings.

## Conclusions

Our results strongly suggest that oral GSH supplementation replenishes the body stores of GSH and significantly reduces oxidative DNA damage in diabetic patients. It also reduces HbA1c within three months and stabilizes it thereafter in diabetic population overall. Elderly sub-group seems to benefit the most as evidenced by a significant decrease in HbA1c and increase in insulin secretion by β-cells over a period of six months. A clinical implication of our study is that oral administration of GSH can be used as an adjunct therapy to anti-diabetic treatment in achieving better glycemic targets, especially in the elderly population.

## Supporting information

Consort checklist

## Data Availability

All datasets generated during and/or analyzed during the current study are not publicly available but are available from the corresponding author on reasonable request.

## Acknowledgements

We thank all patients for their whole-hearted participation in the study. We thank Prof. Satyajit Rath for reading and providing critical comments on the manuscript. We acknowledge the financial help from all the funding agencies mentioned above.

## Authors statements

### Authors’ contributions

SK collected the samples and performed the biochemical assays. JA designed the biochemical assays, supervised the data collection and wrote the manuscript. VG, UD, and SK helped in the recruitment of study participants. AK and PG analyzed the data and performed the statistical analysis. SSG and PG contributed to the design and implementation of the study and editing of the manuscript.

## References

Acharya, J.D., Pande, A.J., Joshi, S.M., Yajnik, C.S., and Ghaskadbi, S.S. 2014. Treatment of hyperglycaemia in newly diagnosed diabetic patients is associated with a reduction in oxidative stress and improvement in *β* -cell function: Glucose Control Reduces Oxidative Stress. Diabetes Metab Res Rev 30(7): 590–598. doi:10.1002/dmrr.2526.

ADA. 2016. Classification and Diagnosis of Diabetes. Diabetes Care 39(Supplement 1): S13– S22. doi:10.2337/dc16-S005.

Allen, J., and Bradley, R.D. 2011. Effects of Oral Glutathione Supplementation on Systemic Oxidative Stress Biomarkers in Human Volunteers. J Altern Complement Med 17(9): 827–833. doi:10.1089/acm.2010.0716.

Ammon, H.P.T., Klumpp, S., Fuý, A., Verspohl, E.J., Jaeschke, H., Wendel, A., and Müller, P. 1989. A possible role of plasma glutathione in glucose-mediated insulin secretion: in vitro and in vivo studies in rats. Diabetologia 32(11). doi:10.1007/BF00264910.

Anjaneyulu, K., Anjaneyulu, R., Sener, A., and Malaisse, W.J. 1982. The stimulus-secretion coupling of glucose-induced insulin release. Thiol: disulfide balance in pancreatic islets. Biochimie 64(1): 29–36. doi:10.1016/S0300-9084(82)80606-8.

Aw, T.Y., Wierzbicka, G., and Jones, D.P. 1991. Oral glutathione increases tissue glutathione in-vivo. Chem Biol Interact 80(1): 89–97. doi:10.1016/0009-2797(91)90033-4.

Baker, M.A., Cerniglia, G.J., and Zaman, A. 1990. Microtiter plate assay for the measurement of glutathione and glutathione disulfide in large numbers of biological samples. Analytical Biochemistry 190(2): 360–365. doi:10.1016/0003-2697(90)90208-Q.

Brownlee, M. 2001. Biochemistry and molecular cell biology of diabetic complications. Nature 414(6865): 813–820. doi:10.1038/414813a.

Brownlee, M. 2005. The Pathobiology of Diabetic Complications: A Unifying Mechanism. Diabetes 54(6): 1615–1625. doi:10.2337/diabetes.54.6.1615.

Bruggeman, B.K., Storo, K.E., Fair, H.M., Wommack, A.J., Carriker, C.R., and Smoliga, J.M. 2019. The absorptive effects of orobuccal non-liposomal nano-sized glutathione on blood glutathione parameters in healthy individuals: A pilot study. PLOS ONE 14(4): e0215815. doi:10.1371/journal.pone.0215815.

Buonocore, D., Grosini, M., Giardina, S., Michelotti, A., Carrabetta, M., Seneci, A., Verri, M., Dossena, M., and Marzatico, F. 2016. Bioavailability Study of an Innovative Orobuccal Formulation of Glutathione. Oxid Med Cell Longev 2016: 3286365. doi:10.1155/2016/3286365.

Cohen, J. 2013. Statistical Power Analysis for the Behavioral Sciences. In 2nd edition. Routledge. doi:10.4324/9780203771587.

De Luca, G., Calpona, P.R., Caponetti, A., Macaione, V., Di Benedetto, A., Cucinotta, D., and Di Giorgio, R.M. 2001. Preliminary report: Amino acid profile in platelets of diabetic patients. Metabolism 50(7): 739–741. doi:10.1053/meta.2001.24193.

DeFronzo, R.A., and Abdul-Ghani, M.A. 2011. Preservation of β-Cell Function: The Key to Diabetes Prevention. J Clin Endocrinol Metab 96(8): 2354–2366. doi:10.1210/jc.2011-0246.

Del Guerra, S., Lupi, R., Marselli, L., Masini, M., Bugliani, M., Sbrana, S., Torri, S., Pollera, M., Boggi, U., Mosca, F., Del Prato, S., and Marchetti, P. 2005. Functional and molecular defects of pancreatic islets in human type 2 diabetes. Diabetes 54(3): 727–735. doi:10.2337/diabetes.54.3.727.

Derubertis, F.R., and Craven, P.A. 1994. Activation of Protein Kinase C in Glomerular Cells in Diabetes: Mechanisms and Potential Links to the Pathogenesis of Diabetic Glomerulopathy. Diabetes 43(1): 1–8. doi:10.2337/diab.43.1.1.

Duckworth, W., Abraira, C., Moritz, T., Reda, D., Emanuele, N., Reaven, P.D., Zieve, F.J., Marks, J., Davis, S.N., Hayward, R., Warren, S.R., Goldman, S., McCarren, M., Vitek, M.E., Henderson, W.G., and Huang, G.D. 2009. Glucose Control and Vascular Complications in Veterans with Type 2 Diabetes. N Engl J Med 360(2): 129–139. doi:10.1056/NEJMoa0808431.

El-Hafidi, M., Franco, M., Ramírez, A.R., Sosa, J.S., Flores, J.A.P., Acosta, O.L., Salgado, M.C., and Cardoso-Saldaña, G. 2018. Glycine Increases Insulin Sensitivity and Glutathione Biosynthesis and Protects against Oxidative Stress in a Model of Sucrose-Induced Insulin Resistance. Oxid Med Cell Longev 2018: 1–12. doi:10.1155/2018/2101562.

Erdennal, M., Sunal, E., and Kanbak, G. 2002. Age-related changes in the glutathione redox system. Cell Biochemistry and Function 20(1): 61–66. doi:10.1002/cbf.937.

Favilli, F., Marraccini, P., Iantomasi, T., and Vincenzini, M.T. 1997. Effect of orally administered glutathione on glutathione levels in some organs of rats: role of specific transporters. Br J Nutr 78(2): 293–300. doi:10.1079/BJN19970147.

Haber, C.A., Lam, T.K.T., Yu, Z., Gupta, N., Goh, T., Bogdanovic, E., Giacca, A., and Fantus, I.G. 2003. N-acetylcysteine and taurine prevent hyperglycemia-induced insulin resistance in vivo: possible role of oxidative stress. Am J Physiol Endocrinol Metab 285(4): E744– E753. doi:10.1152/ajpendo.00355.2002.

Hagen, T.M., and Jones, D.P. 1987. Transepithelial transport of glutathione in vascularly perfused small intestine of rat. Am J Physiol 252(5 Pt 1): G607–613. doi:10.1152/ajpgi.1987.252.5.G607.

Hagen, T.M., Wierzbicka, G.T., Sillau, A.H., Bowman, B.B., and Jones, D.P. 1990. Bioavailability of dietary glutathione: effect on plasma concentration. Am J Physiol 259(4 Pt 1): G524–529. doi:10.1152/ajpgi.1990.259.4.G524.

Hanigan, M.H. 2014. Gamma-Glutamyl Transpeptidase. In Advances in Cancer Research. Elsevier. pp. 103–141. doi:10.1016/B978-0-12-420117-0.00003-7.

Hothorn, T., Hornik, K., van de Wiel, M.A., and Zeileis, A. 2006. A Lego System for Conditional Inference. Am Stat 60(3): 257–263. doi:10.1198/000313006X118430.

Jain, S.K., Velusamy, T., Croad, J.L., Rains, J.L., and Bull, R. 2009. l-Cysteine supplementation lowers blood glucose, glycated hemoglobin, CRP, MCP-1, and oxidative stress and inhibits NF-κB activation in the livers of Zucker diabetic rats. FRBM 46(12): 1633–1638. doi:10.1016/j.freeradbiomed.2009.03.014.

Johansen, J., Harris, A.K., Rychly, D.J., and Ergul, A. 2005. Oxidative stress and the use of antioxidants in diabetes: linking basic science to clinical practice. Cardiovasc Diabetol 4(1): 5. doi:10.1186/1475-2840-4-5.

Kaneto, H., Kajimoto, Y., Miyagawa, J., Matsuoka, T., Fujitani, Y., Umayahara, Y., Hanafusa, T., Matsuzawa, Y., Yamasaki, Y., and Hori, M. 1999. Beneficial effects of antioxidants in diabetes: possible protection of pancreatic beta-cells against glucose toxicity. Diabetes 48(12): 2398–2406. doi:10.2337/diabetes.48.12.2398.

Kannel, W.B., and McGee, D.L. 1979. Diabetes and cardiovascular disease. The Framingham study. JAMA 241(19): 2035–2038. doi:10.1001/jama.241.19.2035.

Kariya, C., Leitner, H., Min, E., van Heeckeren, C., van Heeckeren, A., and Day, B.J. 2007. A role for CFTR in the elevation of glutathione levels in the lung by oral glutathione administration. Am J Physiol Lung Cell Mol Physiol 292(6): L1590–L1597. doi:10.1152/ajplung.00365.2006.

Kolm-Litty, V., Sauer, U., Nerlich, A., Lehmann, R., and Schleicher, E.D. 1998. High glucose-induced transforming growth factor beta1 production is mediated by the hexosamine pathway in porcine glomerular mesangial cells. J Clin Invest 101(1): 160–169. doi:10.1172/JCI119875.

Kovacs-Nolan, J., Rupa, P., Matsui, T., Tanaka, M., Konishi, T., Sauchi, Y., Sato, K., Ono, S., and Mine, Y. 2014. In Vitro and ex Vivo Uptake of Glutathione (GSH) across the Intestinal Epithelium and Fate of Oral GSH after in Vivo Supplementation. J Agric Food Chem 62(39): 9499–9506. doi:10.1021/jf503257w.

Kulkarni, R., Acharya, J., Ghaskadbi, S., and Goel, P. 2014. Thresholds of Oxidative Stress in Newly Diagnosed Diabetic Patients on Intensive Glucose-Control Therapy. PLoS ONE 9(6): e100897. doi:10.1371/journal.pone.0100897.

Lang, C.A., Naryshkin, S., Schneider, D.L., Mills, B.J., and Lindeman, R.D. 1992. Low blood glutathione levels in healthy aging adults. J Lab Clin Med 120(5): 720–725.

Lee, A.Y.W., and Chung, S.S.M. 1999. Contributions of polyol pathway to oxidative stress in diabetic cataract. FASEBj. 13(1): 23–30. doi:10.1096/fasebj.13.1.23.

Lenzen, S., Drinkgern, J., and Tiedge, M. 1996. Low antioxidant enzyme gene expression in pancreatic islets compared with various other mouse tissues. FRBM 20(3): 463–466. doi:10.1016/0891-5849(96)02051-5.

Martín-Gallán, P., Carrascosa, A., Gussinyé, M., and Domínguez, C. 2003. Biomarkers of diabetes-associated oxidative stress and antioxidant status in young diabetic patients with or without subclinical complications. FRBM 34(12): 1563–1574. doi:10.1016/S0891-5849(03)00185-0.

Matsubara, L.S., and Machado, P.E. 1991. Age-related changes of glutathione content, glutathione reductase and glutathione peroxidase activity of human erythrocytes. Braz J Med Biol Res 24(5): 449–454.

Modak, M.A., Parab, P.B., and Ghaskadbi, S.S. 2009. Pancreatic Islets Are Very Poor in Rectifying Oxidative DNA Damage: Pancreas 38(1): 23–29. doi:10.1097/MPA.0b013e318181da4e.

Modak, M.A., Parab, P.B., and Ghaskadbi, S.S. 2014. Tissue specific oxidative stress profile in relation to glycaemic regulation in mice. Diabetes Metab Res Rev 30(1): 31–41. doi:10.1002/dmrr.2460.

Moini, H., Tirosh, O., Park, Y.C., Cho, K.-J., and Packer, L. 2002. R-α-Lipoic Acid Action on Cell Redox Status, the Insulin Receptor, and Glucose Uptake in 3T3-L1 Adipocytes. Arch Biochem Biophys 397(2): 384–391. doi:10.1006/abbi.2001.2680.

Njälsson, R., and Norgren, S. 2007. Physiological and pathological aspects of GSH metabolism: Disorders of glutathione metabolism. Acta Paediatrica 94(2): 132–137. doi:10.1111/j.1651-2227.2005.tb01878.x.

Nuttall, S., Martin, U., Sinclair, A., and Kendall, M. 1998. Glutathione: in sickness and in health. The Lancet 351(9103): 645–646. doi:10.1016/S0140-6736(05)78428-2.

Page, K.A., and Reisman, T. 2013. Interventions to preserve beta-cell function in the management and prevention of type 2 diabetes. Curr Diab Rep 13(2): 252–260. doi:10.1007/s11892-013-0363-2.

Paolisso, G., Di Maro, G., Pizza, G., D’Amore, A., Sgambato, S., Tesauro, P., Varricchio, M., and D’Onofrio, F. 1992a. Plasma GSH/GSSG affects glucose homeostasis in healthy subjects and non-insulin-dependent diabetics. Am J Physiol Endocrinol Metab 263(3): E435–E440. doi:10.1152/ajpendo.1992.263.3.E435.

Paolisso, G., Giugliano, D., Pizza, G., Gambardella, A., Tesauro, P., Varricchio, M., and D’Onofrio, F. 1992b. Glutathione Infusion Potentiates Glucose-Induced Insulin Secretion in Aged Patients With Impaired Glucose Tolerance. Diabetes Care 15(1): 1–7. doi:10.2337/diacare.15.1.1.

Picu, A., Petcu, L., Ştefan, S., Mitu, M., Lixandru, D., Ionescu-Tîrgovişte, C., Pîrcălăbioru, G.G., Ciulu-Costinescu, F., Bubulica, M.-V., and Chifiriuc, M.C. 2017. Markers of Oxidative Stress and Antioxidant Defense in Romanian Patients with Type 2 Diabetes Mellitus and Obesity. Molecules 22(5): 714. doi:10.3390/molecules22050714.

Richie, J.P., Nichenametla, S., Neidig, W., Calcagnotto, A., Haley, J.S., Schell, T.D., and Muscat, J.E. 2015. Randomized controlled trial of oral glutathione supplementation on body stores of glutathione. Eur J Nutr 54(2): 251–263. doi:10.1007/s00394-014-0706-z.

Sawilowsky, S.S. 2003. A Different Future For Social And Behavioral Science Research. J Mod Appl Stat Methods 2(1): 128–132. doi:10.22237/jmasm/1051747860.

Schmitt, B., Vicenzi, M., Garrel, C., and Denis, F.M. 2015. Effects of N-acetylcysteine, oral glutathione (GSH) and a novel sublingual form of GSH on oxidative stress markers: A comparative crossover study. Redox Biology 6: 198–205. doi:10.1016/j.redox.2015.07.012.

Sekhar, R.V., McKay, S.V., Patel, S.G., Guthikonda, A.P., Reddy, V.T., Balasubramanyam, A., and Jahoor, F. 2011a. Glutathione Synthesis Is Diminished in Patients With Uncontrolled Diabetes and Restored by Dietary Supplementation With Cysteine and Glycine. Diabetes Care 34(1): 162–167. doi:10.2337/dc10-1006.

Sekhar, R.V., Patel, S.G., Guthikonda, A.P., Reid, M., Balasubramanyam, A., Taffet, G.E., and Jahoor, F. 2011b. Deficient synthesis of glutathione underlies oxidative stress in aging and can be corrected by dietary cysteine and glycine supplementation. Am J Clin Nutr 94(3): 847–853. doi:10.3945/ajcn.110.003483.

Shinohara, M., Thornalley, P.J., Giardino, I., Beisswenger, P., Thorpe, S.R., Onorato, J., and Brownlee, M. 1998. Overexpression of glyoxalase-I in bovine endothelial cells inhibits intracellular advanced glycation end product formation and prevents hyperglycemia-induced increases in macromolecular endocytosis. J Clin Invest 101(5): 1142–1147. doi:10.1172/JCI119885.

Sinha, R., Sinha, I., Calcagnotto, A., Trushin, N., Haley, J.S., Schell, T.D., and Richie, J.P. 2018. Oral supplementation with liposomal glutathione elevates body stores of glutathione and markers of immune function. Eur J Clin Nutr 72(1): 105–111. doi:10.1038/ejcn.2017.132.

Song, F., Jia, W., Yao, Y., Hu, Y., Lei, L., Lin, J., Sun, X., and Liu, L. 2007. Oxidative stress, antioxidant status and DNA damage in patients with impaired glucose regulation and newly diagnosed Type 2 diabetes. Clinical Science 112(12): 599–606. doi:10.1042/CS20060323.

Tiedge, M., Lortz, S., Drinkgern, J., and Lenzen, S. 1997. Relation between antioxidant enzyme gene expression and antioxidative defense status of insulin-producing cells. Diabetes 46(11): 1733–1742. doi:10.2337/diab.46.11.1733.

Townsend, D.M., Tew, K.D., and Tapiero, H. 2003. The importance of glutathione in human disease. Biomed Pharmacother 57(3–4): 145–155. doi:10.1016/S0753-3322(03)00043-X.

Ueno, Y., Kizaki, M., Nakagiri, R., Kamiya, T., Sumi, H., and Osawa, T. 2002. Dietary Glutathione Protects Rats from Diabetic Nephropathy and Neuropathy. The Journal of Nutrition 132(5): 897–900. doi:10.1093/jn/132.5.897.

Zhang, J., An, H., Ni, K., Chen, B., Li, H., Li, Y., Sheng, G., Zhou, C., Xie, M., Chen, S., Zhou, T., Yang, G., Chen, X., Wu, G., Jin, S., and Li, M. 2019. Glutathione prevents chronic oscillating glucose intake-induced β-cell dedifferentiation and failure. Cell Death & Disease 10(4). doi:10.1038/s41419-019-1552-y.

