## Supplementary figures for "Randomised clinical trial of long term glutathione supplementation offers protection from oxidative damage, improves HbA1c in elderly type 2 diabetic patients"

**Fig. S1**

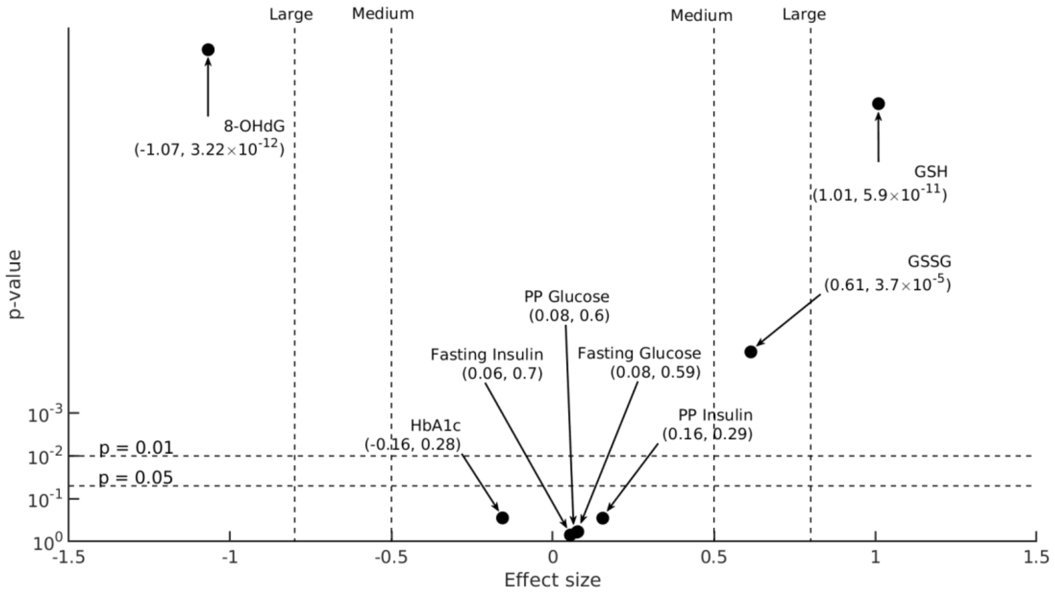


**Fig. S1 The effect size of changes in blood biochemical parameters** 6-month changes in biochemical parameters (X= H, FPG, FPI, PPG, PPI, GSH, GSSG, 8-OHdG) from $\alpha$ to $\gamma$visit in D and DG groups, $\Delta X_{\gamma-\alpha}^{D}$and $\Delta X_{\gamma-\alpha}^{DG}$respectively (sample sizes for HbA1c, FPG, FPI are:$N_D$= 90, $N_{DG}$= 104, for PPG and PPI:$N_D$= 90, $N_{DG}$= 102, and for GSH, GSSG:$N_D$= 89, $N_{DG}$= 101, for 8-OHdG:$N_D$= 91, $N_{DG}$= 104), are compared here on a statistical significance (y-axis) versus effect size (x-axis) plot. Effect size (Cohen's d) computed between 6-month changes in the concentration of biochemical variables, X, in D and DG group ($\Delta X_{\gamma-\alpha}^{D}$and $\Delta X_{\gamma-\alpha}^{DG}$) are denoted on the x-axis. The p-value of comparison between the group-wise means of 6-month changes in X's concentration, $\Delta\bar{X}_{\gamma-\alpha}^{D}$and $\Delta\bar{X}_{\gamma-\alpha}^{DG}$, obtained using two-sample, two-sided t-tests are displayed on the y-axis. Two horizontal dotted lines denote 95% and 99% significance levels. Effect size takes either a positive or negative sign based on the direction of change: a positive effect size increases towards the right and a negative effect towards the left. Vertical dotted lines represent "Medium" and "Large" effects at 0.5 and 0.8, respectively.

**Fig. S2**

a

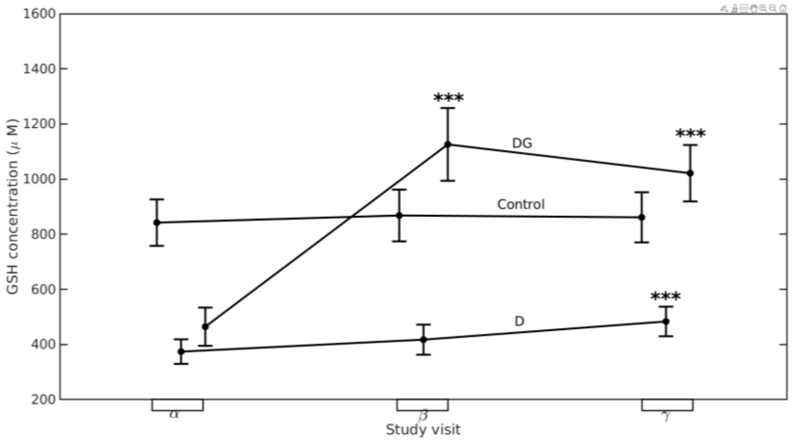


b

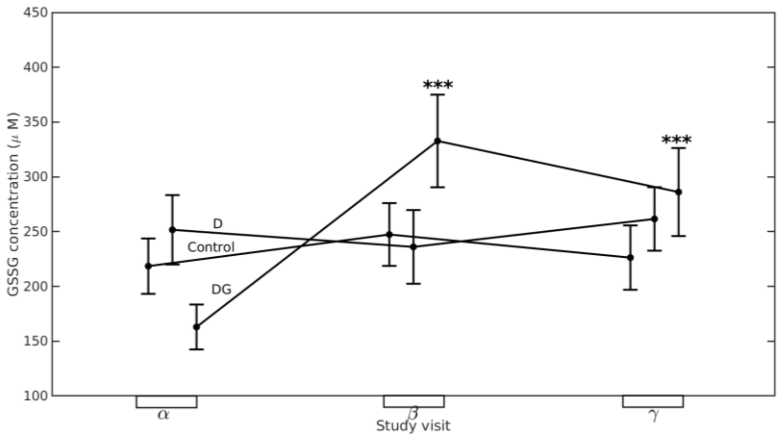


**Fig S2. Longitudinal changes in the concentration of (a) GSH and (b) GSSG.** Mean (black dots) and 95% confidence interval (whiskers) of (a) GSH and (b) GSSG concentrations at $\alpha$ visit ($N_C = 104, N_D = 100, N_{DG} = 101$), $\beta$ visit ($N_C = 104, N_D = 100, N_{DG} = 101$), and $\boldsymbol{\gamma}$ visit ($N_C = 104, N_D = 89, N_{DG} = 101$) are shown here for Control, D, and DG groups. Significance levels (*) displayed above $\beta$, and $\boldsymbol{\gamma}$ visits denote the comparisons with $\alpha$ visit using paired sample t-tests. Significance levels are *p < 0.05, **p < 0.01, ***p < 0.001 for respective comparisons.

**Fig.S3**

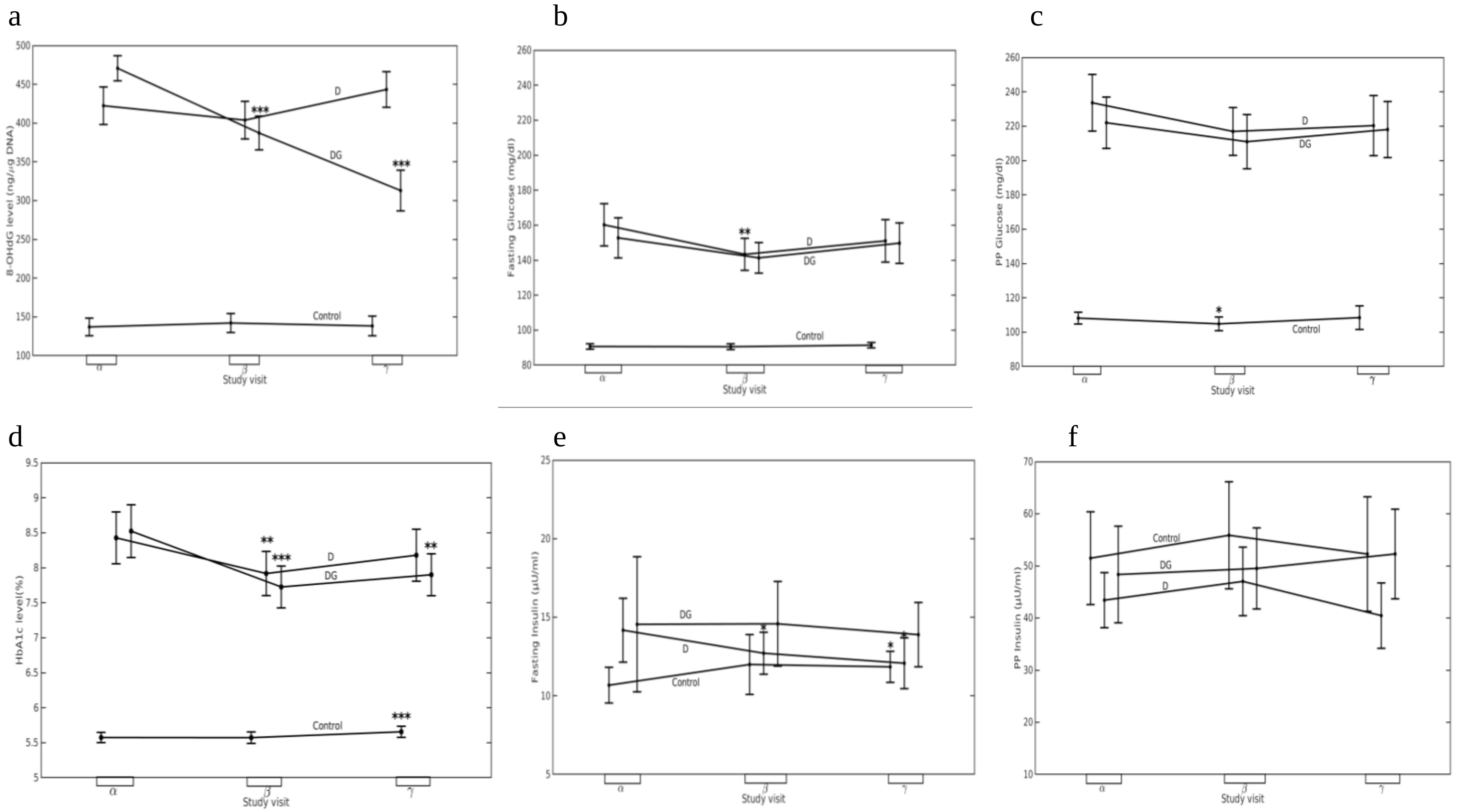


**Fig. S3 Effect of oral GSH supplementation on blood glycaemic parameters.** Mean (black dots) and 95% confidence intervals (whiskers) of (a) 8-OHdG, (b) FPG, (c) PPG, (d) HbA1c, (e) FPI and (f) PPI at $\alpha$ visit (sample sizes for 8-OHdG $N_C = 104$, $N_D$=91, $N_{DG}$=104), β ($N_C = 104, N_D$=91, $N_{DG}$=104), and γ visit ($N_C = 104$, $N_D$=91, $N_{DG}$=104, for HbA1c, FPG, FPI, PPG are $N_C = 104$, $N_D$=102, $N_{DG}$=104, for PPI:$N_C = 103, N_D$=102, $N_{DG}$=104, and for GSH, GSSG:$N_C = 104, N_D$=100, $N_{DG}$=101), $\beta$ visit (for HbA1c, FPG, FPI :$N_C = 104 N_D$=102, $N_{DG}$=103, for PPG:$N_C = 103, N_D$=102, $N_{DG}$=102, for PPI:$N_C = 103, N_D$=101, $N_{DG}$=102, and for GSH, GSSG:$N_C = 104, N_D$=100, $N_{DG}$=101), and $\gamma$ visit (for HbA1c, FPG, and FPI:$N_C = 104$, $N_D$=90, $N_{DG}$=104, for PPG, PPI:$N_C = 104$ , $N_D$=90, $N_{DG}$=102, and for GSH, GSSG:$N_C = 104, N_D$=89, $N_{DG}$=101) are shown here for Control, D, and DG groups. Significance levels (*) displayed above $\beta$ and $\boldsymbol{\gamma}$ visits denote the comparisons with $\alpha$visit using paired sample t-tests. Significance levels are *p < 0.

**Fig. S4**

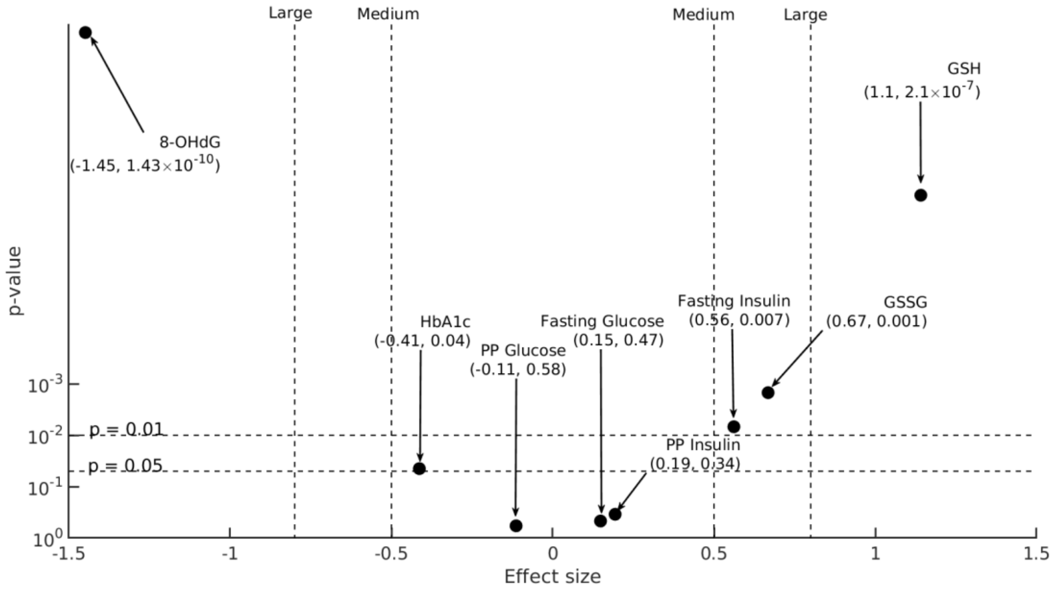


**Figure S4. The effect size of changes in blood biochemical parameters in elderly diabetic patients.** 6-month changes in biochemical parameters (X= H, FPG, FPI, PPG, PPI, GSH, GSSG, 8-OHdG) from $\alpha$ to $\gamma$visit in elderly (age > 55 years old) D and DG group subjects, $\Delta X^{D}_{\gamma-\alpha}$and $\Delta X^{DG}_{\gamma-\alpha}$respectively (sample sizes for HbA1c, FPG, FPI are:$N_D$= 44, $N_{DG}$= 54, and for PPG, PPI, GSH, GSSG are:$N_D$= 44, $N_{DG}$= 53, , for 8-OHdG:$N_D$= 45, $N_{DG}$= 54), are compared here on a statistical significance (y-axis) versus effect size (x-axis) plot. Effect size (Cohen's d) calculated between the 6-month changes in X's concentration in the subgroups of D and DG ($\Delta X^{D}_{\gamma-\alpha}$and $\Delta X^{DG}_{\gamma-\alpha}$respectively) are displayed on the x-axis. The p-value of comparison between the subgroup-wise means of 6-month changes in X's concentration, $\Delta\bar{X}^{D}_{\gamma-\alpha}$ and $\Delta\bar{X}^{DG}_{\gamma-\alpha}$, obtained using two-sample, two-sided t-tests. Effect size takes either a positive or negative sign based on the direction of change: a positive effect size increases towards the right and a negative effect towards the left. Vertical dotted lines represent "Medium" and "Large" effects at 0.5 and 0.8, respectively.

**Fig. S5**

a b

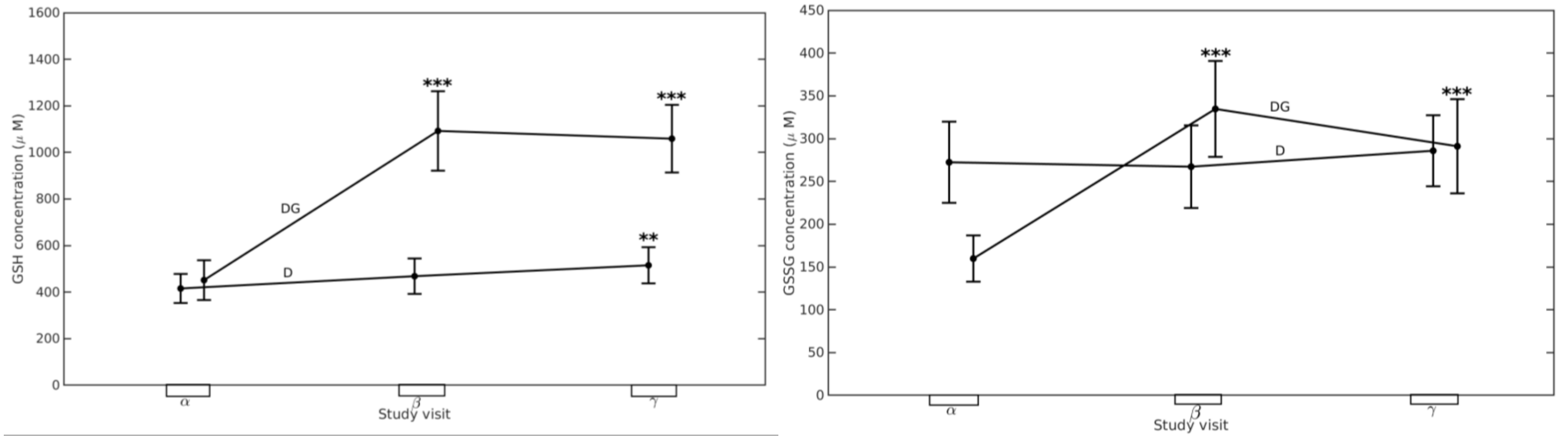


**Fig. S5 Serial changes in the concentration of (a) GSH and (b) GSSG in elderly diabetic subjects.** Mean (black dots) and 95% confidence interval (whiskers) of (a) GSH and (b) GSSG concentrations in elderly subjects of D and DG groups at $\alpha$ visit ($N_D = 50, N_{DG} = 53$), $\beta$ visit ($N_D = 50, N_{DG} = 53$), and $\boldsymbol{\gamma}$ visit ($N_D = 44, N_{DG} = 53$) are shown here. Significance levels (*) displayed above $\beta$, and $\boldsymbol{\gamma}$ visits denote the comparisons with $\alpha$ visit using permutation tests. Significance levels are *p < 0.05, **p < 0.01, ***p < 0.001 for respective comparisons (See Fig. S7 for alternative comparisons using paired sample t-tests instead). Confidence interval whiskers are not displayed for the control group since there were only seven elderly subjects in that group.

**Fig. S6**

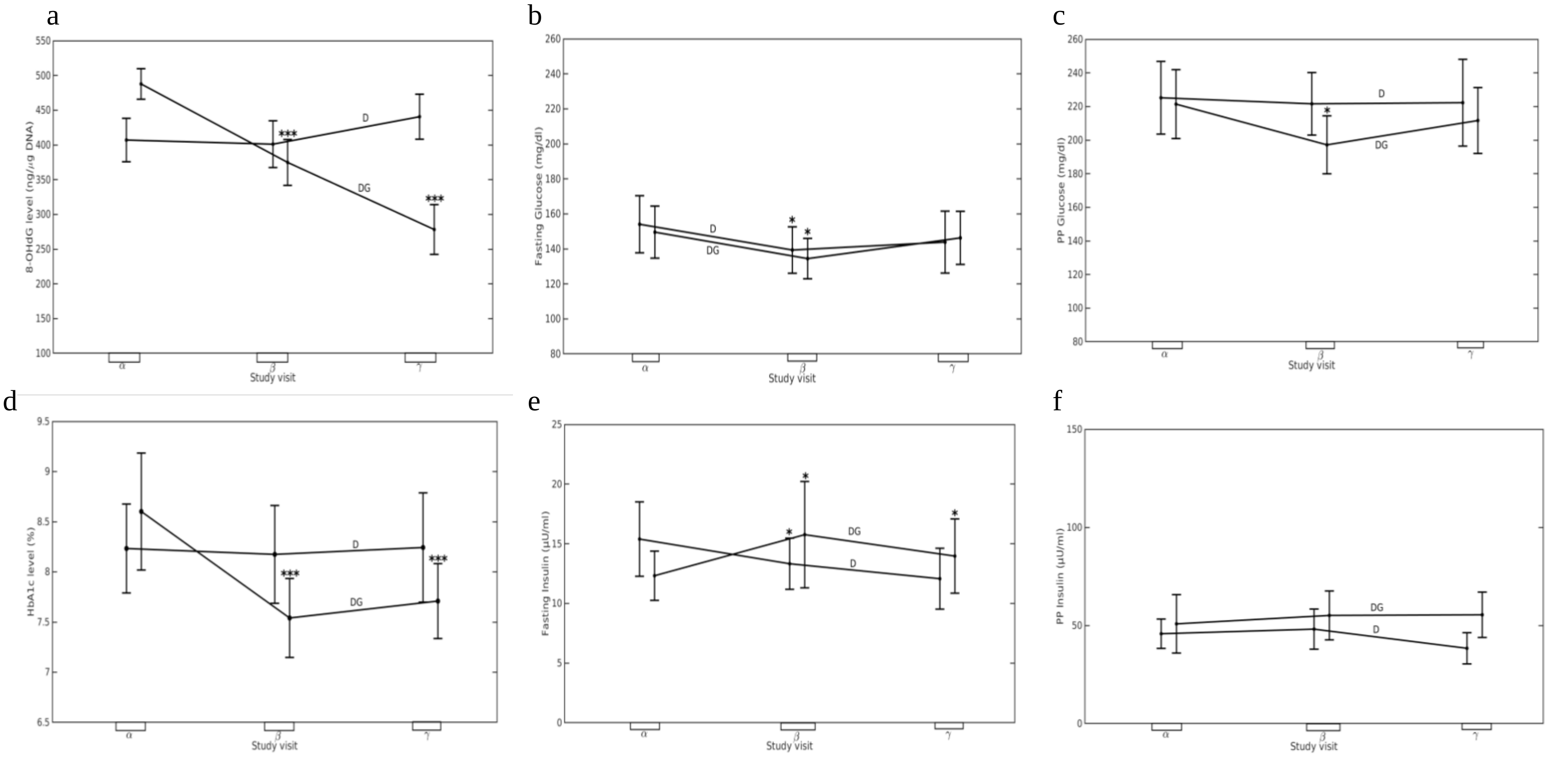


**Fig. S6 Longitudinal changes in the concentration of biochemical parameters in elderly diabetic subjects.** Mean (black dots) and 95% confidence interval (whiskers) of (a) 8-OHdG, (b) FPG, (c) PPG, (d) HbA1c, (e) FPI and (f) PPI concentrations in elderly subjects of D and DG groups at $\alpha$ visit (sample sizes : $N_D = 51, N_{DG} = 54$), $\beta$visit (sample sizes for 8-OHdG, FPG, FPI, HbA1c: $N_D = 51, N_{DG} = 54$, for PPG: $N_D = 51, N_{DG} = 53$,and for PPI:$N_D = 50, N_{DG} = 53$), and $\boldsymbol{\gamma}$ visit (for 8-OHdG $N_D = 45, N_{DG} = 54$, for FPG, FPI, HbA1c: $N_D = 44, N_{DG} = 54$, and for PPG, PPI: $N_D = 44, N_{DG} = 53$) are shown here. Significance levels (*) displayed above $\beta$ and $\boldsymbol{\gamma}$ visits denote the comparisons with $\alpha$ visit using two-sample permutation tests. Significance levels are *p < 0.05, **p < 0.01, ***p < 0.001 for respective comparisons (See Fig. S8 for alternative comparisons using paired sample t-tests instead). Confidence interval whiskers are not displayed for the control since there were only seven elderly subjects in that group.

**Fig. S7**

a b

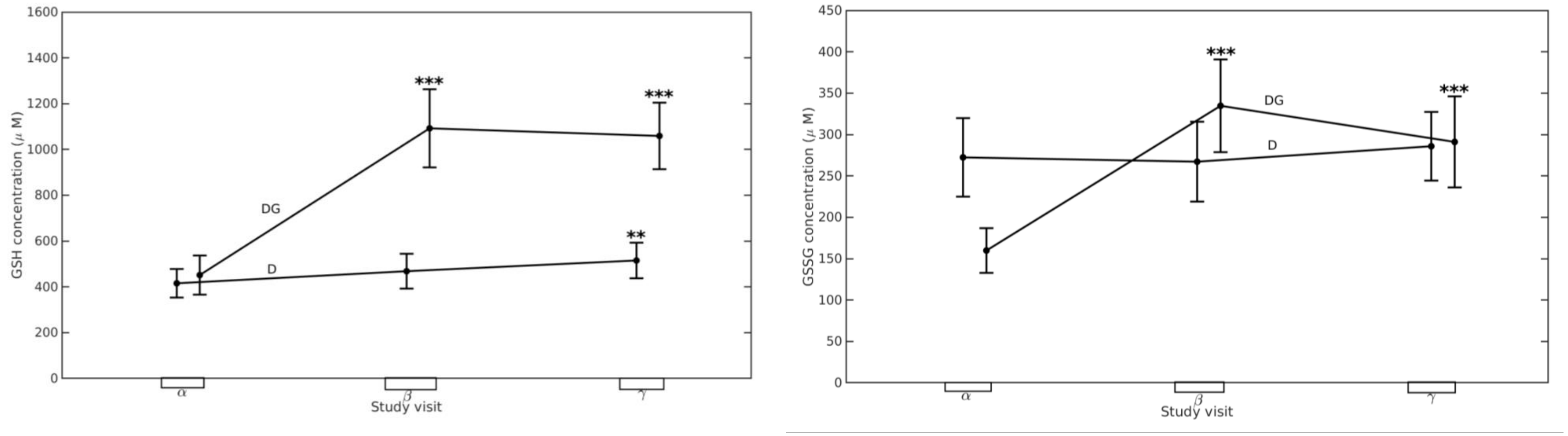


**Fig. S7 Serial changes in the concentration of (a) GSH and (b) GSSG in elderly diabetic subjects.** Mean (black dots) and 95% confidence interval (whiskers) of (a) GSH and (b) GSSG concentrations in elderly subjects of D and DG groups at $\alpha$ visit ($N_D = 50, N_{DG} = 53$), $\beta$ visit ($N_D = 50, N_{DG} = 53$), and $\boldsymbol{\gamma}$ visit ($N_D = 44, N_{DG} = 53$) are shown here. Significance levels (*) displayed above $\beta$, and $\boldsymbol{\gamma}$ visits denote the comparisons with $\alpha$visit using paired t-tests. Significance levels are *p < 0.05, **p < 0.01, ***p < 0.001 for respective comparisons. Confidence interval whiskers are not displayed for the control group since there were only seven elderly subjects in that group.

**Fig. S8**

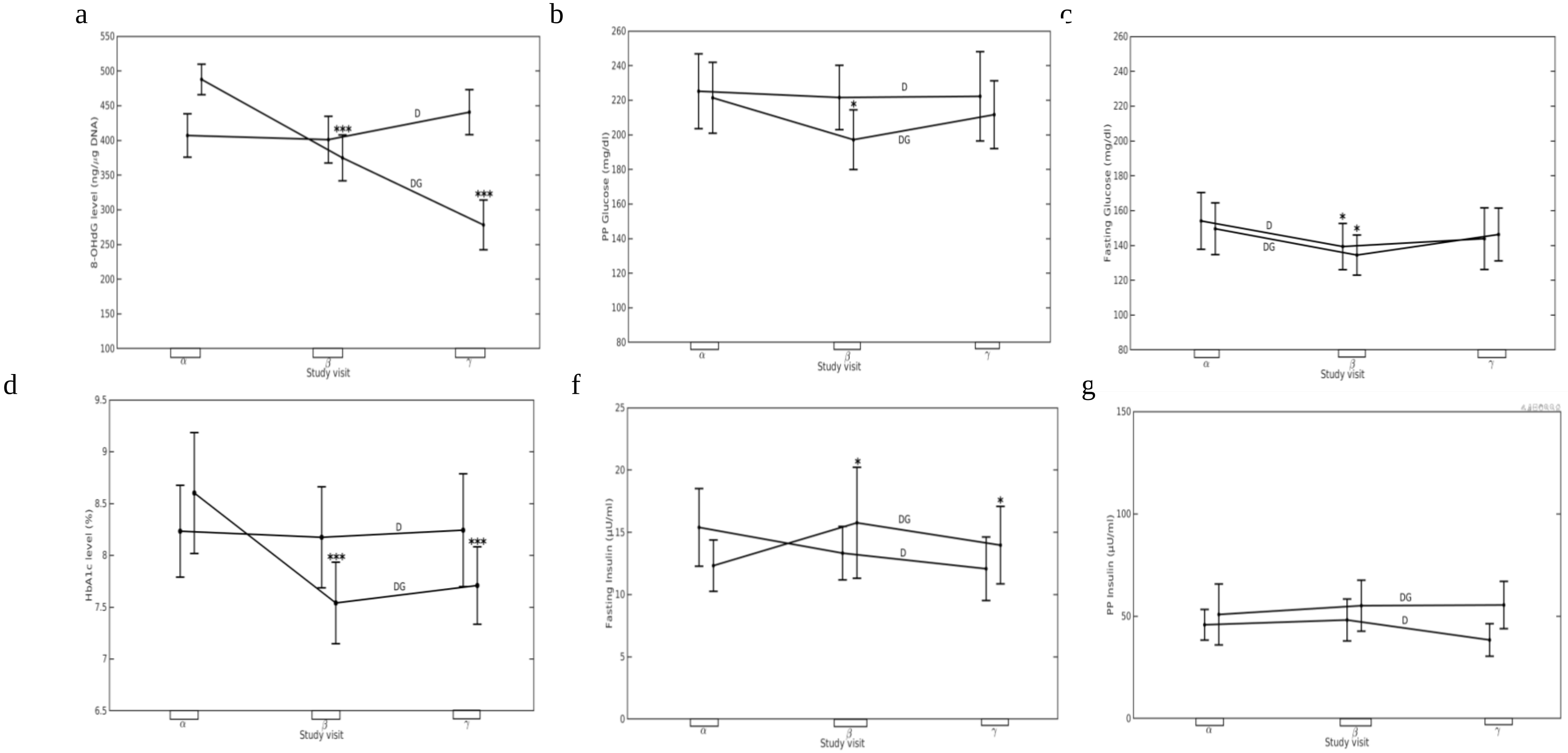


**Fig. S8 Effect of oral GSH supplementation on blood glycaemic parameters in elderly diabetic subjects.** Mean (black dots) and 95% confidence interval (whiskers) of a) 8-OHdG, (b) FPG, (c) PPG, (d) HbA1c, (e) FPI and (f) PPI concentrations in elderly subjects of D and DG groups at $\alpha$ visit (sample sizes: $N_D = 51, N_{DG} = 54$), $\beta$ visit (sample sizes for 8-OHdG, FPG, FPI, HbA1c: $N_D = 51, N_{DG} = 54$, for PPG: $N_D = 51, N_{DG} = 53$,and for PPI:$N_D = 50, N_{DG} = 53$), and $\boldsymbol{\gamma}$ visit (for 8-OHdG $N_D = 45, N_{DG} = 54$, for FPG, FPI, HbA1c: $N_D = 44, N_{DG} = 54$, and for PPG and PPI: $N_D = 44, N_{DG} = 53$) are shown here. Significance levels (*) displayed above $\beta$,and $\boldsymbol{\gamma}$ visits denote the comparisons with $\alpha$visit using paired sample t-tests. Significance levels are *p < 0.05, **p < 0.01, ***p < 0.001 for respective comparisons. Confidence interval whiskers are not displayed for the control group since there were only seven elderly subjects in that group.
