## Supplementary tables for "Randomised clinical trial of long term glutathione supplementation offers protection from oxidative damage, improves HbA1c in elderly type 2 diabetic patients"

**Table S1.**

a

| **Sr No.** | **Type of treatment in Diabetic (D) group** | **No. of subjects** |
|---|---|---|
| 1 | Biguanides | 19 |
| 2 | Sulphonylureas | 10 |
| 3 | Thiazolidinediones | 1 |
| 4 | Meglitinides | 1 |
| 5 | Biguanides & Sulphonylureas | 35 |
| 6 | Biguanides & DPP-4 inhibitor | 6 |
| 7 | Biguanides & Insulin | 2 |
| 8 | Sulphonylureas & glucosidases inhibitor | 1 |
| 9 | Biguanides & DPP-4 inhibitor | 4 |
| 11 | DPP-4 inhibitor & Sulphonylureas | 1 |
| 12 | Biguanides & Sulphonylureas &Thiazolidinediones | 2 |
| 13 | Biguanides & Sulphonylureas & DPP-4 inhibitor | 2 |
| 14 | Biguanides & Sulphonylureas & Insulin | 1 |
| 15 | Biguanides & Sulphonylureas & α-glucosidases inhibitor | 5 |
| 16 | Biguanides & DPP-4 inhibitor & α-glucosidases inhibitor | 1 |
| 17 | Controlled by exercise | 10 |
| | Total | 101 |

b

| **Sr No.** | **Type of treatment Diabetic with Glutathione (DG) group** | **No. of subjects** |
|---|---|---|
| 1 | Biguanides | 35 |
| 2 | Sulphonylureas | 2 |
| 3 | Insulin | 1 |
| 4 | Biguanides & Sulphonylureas | 26 |
| 5 | Biguanides & DPP-4 inhibitor | 7 |
| 6 | Biguanides & Insulin | 1 |
| 7 | Biguanides & $\alpha$-glucosidases inhibitor | 1 |
| 8 | DPP-4 inhibitor & $\alpha$-glucosidases inhibitor | 1 |
| 9 | DPP-4 inhibitor & Insulin | 1 |
| 10 | Biguanides & Sulphonylureas &Thiazolidinediones | 1 |
| 11 | Biguanides & Sulphonylureas & DPP-4 inhibitor | 11 |
| 12 | Biguanides & Sulphonylureas & Insulin | 1 |
| 13 | Biguanides & Sulphonylureas & $\alpha$-glucosidases inhibitor | 1 |
| 14 | Biguanides & $\alpha$-glucosidases inhibitor & DPP-4 inhibitor | 1 |
| 15 | Controlled by exercise | 5 |
| | Total | 95 |

**Table S1. Anti-diabetic treatment in D (A) and DG (B) groups.** Number of subjects in D and DG groups with different types of anti-diabetic treatment are listed here.

**Table S2.**

| Groups | Variables | $\alpha$ Visit<br>--------<br>Mean (95% CI) | $\beta$ Visit<br>--------<br>Mean (95% CI) | $\gamma$ Visit<br>--------<br>Mean (95% CI) |
|---|---|---|---|---|
| **Control** | **HbA1c** (%) | 5.6 (5.5-5.6) | 5.6 (5.5-5.7) | 5.7 (5.6-5.7)<br>*** |
| | **FPG** (mg/dl) | 91 (89-92) | 91 (89-92) | 91 (90-93) |
| | **PPG** (mg/dl) | 108 (105-112) | 105 (101-109)<br>* | 108 (102-115) |
| | **FPI** (µU/ml) | 10.7 (9.5-11.8) | 12 (10.1-13.9) | 11.8 (10.9-12.8)<br>* |
| | **PPI** (µU/ml) | 51.5 (42.6-60.4) | 55.9 (45.6-66.2) | 52.3 (41.3-63.3) |
| | **GSH** (µM) | 842 (758-926) | 868 (774-962) | 861 (770-952) |
| | GSSG (µM) | 218 (193-244) | 247 (219-276) | 226 (197-256) |
| | **8-OHdG** (ng/µg DNA) | 137.03 (125.66-148.31) | 42.09 (129.9-154.28) | 129.9 (154.28-138.31) |
| **D** | **HbA1c** (%) | 8.4 (8.1-8.8) | 7.9 (7.6-8.2)<br>** | 8.2 (7.8-8.6) |
| | **FPG** (mg/dl) | 160 (148-172) | 143 (134-152)<br>** | 151 (139-163) |
| | **PPG** (mg/dl) | 14.2 (12.1-16.2) | 12.7 (11.4-14.0)<br>* | 12.1 (10.5-13.7)<br>* |
| | **FPI** (µU/ml) | 234 (217-250) | 217 (203-231) | 220 (203-238) |
| | **PPI** (µU/ml) | 43.4 (38.1-48.7) | 47 (40.5-53.6) | 40.5 (34.2-46.7) |
| | **GSH** (µM) | 395 (351-440) | 428 (376-480) | 484 (430-537)<br>*** |

| | | | | |
|---|---|---|---|---|
| | **GSSG** (µM) | 249 (220-279) | 236 (205-268) | 262 (233-291) |
| | **8-OHdG** (ng/µg DNA) | 422.41 (398.16-446.66) | 403.82 (379.54-428.09) | 443.32 (420.32-466.31) |
| **DG** | **HbA1c** (%) | 8.5 (8.1-8.9) | 7.7 (7.4-8) *** | 7.9 (7.6-8.2) *** |
| | **FPG** (mg/dl) | 153 (141-164) | 141 (133-150) | 150 (138-161) |
| | **PPG** (mg/dl) | 14.6 (10.2-18.9) | 14.6 (11.9-17.3) | 13.9 (11.8-15.9) |
| | **FPI** (µU/ml) | 222 (207-237) | 211 (195-227) | 218 (202-234) |
| | **PPI** (µU/ml) | 48.4 (39.1-57.6) | 49.5 (41.7-57.3) | 52.3 (43.7-60.9) |
| | **GSH** (µM) | 465 (395-534) | 1126 (994-1258) *** | 1022 (919-1124) *** |
| | **GSSG** (µM) | 163 (142-183) | 333 (290-375) *** | 286 (246-326) *** |
| | **8-OHdG** (ng/µg DNA) | 470.7(454.48-486.93) | 387.21 (365.44-408.98) *** | 312.96 (286.7-339.22) *** |

**Table S2. Biochemical measurements at different visits in Control, D and DG groups.** Biochemical measurements of subjects in Control, D, and DG groups at $\alpha$, β, and $\gamma$visits are given here with mean and 95% confidence interval. Significance levels (*) displayed at $\beta$,and $\gamma$visits denote the comparisons with $\alpha$visit using permutation tests. Significance levels are *p < 0.05, **p < 0.01, ***p < 0.001 for respective comparisons (See Table S6 for p-values of these comparisons using permutation tests and alternate comparisons using t-tests).

**Table S3.**

| Variables | Control versus D (p-value) | | Control versus DG (p-value) | | D versus DG (p-value) | |
|---|---|---|---|---|---|---|
| | Permutation test | t-test | Permutation test | t-test | Permutation test | t-test |
| **Age** | $1.41\times 10^{-16}$ *** | $3.6\times 10^{-17}$ *** | $7.96\times 10^{-18}$ *** | $2.12\times 10^{-18}$ *** | 0.77 | 0.77 |
| **BMI** | 0.88 | 0.88 | 0.10 | 0.10 | 0.071 | 0.073 |
| **HbA1c** | $4.12\times 10^{-46}$ *** | $3.79\times 10^{-35}$ *** | $8.41\times 10^{-55}$ *** | $1.36\times 10^{-35}$ *** | 0.72 | 0.72 |
| **FPG** | $2.89\times 10^{-36}$ *** | $9.92\times 10^{-36}$ *** | $8.18\times 10^{-32}$ *** | $2.26\times 10^{-21}$ *** | 0.38 | 0.38 |
| **FPI** | 0.002** | 0.003 ** | 0.029* | 0.086 | 0.93 | 0.88 |
| **PPG** | $1.9\times 10^{-44}$ *** | $2.23\times 10^{-34}$ *** | $4.32\times 10^{-45}$ *** | $6.5\times 10^{-34}$ *** | 0.302 | 0.30 |
| **PPI** | 0.13 | 0.13 | 0.63 | 0.63 | 0.37 | 0.36 |
| **GSH** | $7.51\times 10^{-19}$ *** | $4.94\times 10^{-17}$ *** | $3.57\times 10^{-11}$ *** | $9.23\times 10^{-11}$ *** | 0.098 | 0.098 |
| **GSSG** | 0.12 | 0.12 | $8.33\times 10^{-4}$ *** | $8.80\times 10^{-4}$*** | $2.99\times 10^{-6}$ *** | $3.7\times 10^{-6}$ *** |
| **8-OHdG** | $2.94\times 10^{-46}$ *** | $1.17\times 10^{-53}$ *** | $5.46\times 10^{-58}$ *** | $4.46\times 10^{-85}$ *** | 0.0012 ** | 0.0011 ** |

**Table S3. Inter-group comparisons of baseline characteristics.** Variables at the α visit in different groups were compared using permutation tests and t-tests. p-values of comparison are shown here in this table. Significance levels are *$p < 0.05$, **$p < 0.01$, ***$p < 0.001$ for respective comparisons.

**Table S4.**

| **Biochemical variables (X)** | **D group** | | **DG group** | |
|---|---|---|---|---|
| | $\Delta\bar{X}^{D}_{\gamma-\alpha}$ **(**$s_1$**)** | $N_D$ | $\Delta\bar{X}^{DG}_{\gamma-\alpha}$ **(**$s_2$**)** | $N_{DG}$ |
| **HbA1c** (%) | -0.3 (1.9) | 90 | -0.6 (1.9) | 104 |
| **FPG** (mg/dl) | -9 (68) | 90 | -3 (72) | 104 |
| **PPG** (mg/dl) | -13 (104) | 90 | -5 (96) | 102 |
| **FPI** (µU/ml) | -1.6(6.9) | 90 | -0.7 (21.6) | 104 |
| **PPI** (µU/ml) | -1.3 (30.4) | 90 | 4.8 (45) | 102 |
| **GSH** (µM) | 109 (274) | 89 | 557 (550) | 101 |
| **GSSG** (µM) | 10 (125) | 89 | 123 (224) | 101 |
| **8-OHdG (ng/µg DNA)** | 24.25 (170.11) | 91 | -157.74 (170.99) | 104 |

**Table S4. Six-month biochemical changes in D and DG groups.** Mean ($\Delta\bar{X}^{D}_{\gamma-\alpha}$ and $\Delta\bar{X}^{DG}_{\gamma-\alpha}$) and standard deviation ($s_1$ and $s_2$) of 6-month biochemical changes in D, and DG groups are given here in separate columns, where $N_D$ and $N_{DG}$ are the number of individuals in D and DG, respectively.

**Table S5.**

| Groups | Variables | $\alpha$Visit | β Visit | $\gamma$ Visit |
|---|---|---|---|---|
| | | Mean (95% CI) | Mean (95% CI) | Mean (95% CI) |
| **D** | **HbA1c** (%) | 8.2 (7.8-8.7) | 8.2 (7.7-8.7) | 8.2 (7.7-8.8) |
| | **FPG** (mg/dl) | 154 (138-170) | 139 (126-153) * | 144 (126-162) |
| | **FPI** (µU/ml) | 15.4 (12.3-18.5) | 13.3 (11.2-15.5) * | 12.1 (9.5-14.6)* |
| | **PPG** (mg/dl) | 225 (204-247) | 222 (203-240) | 222 (196-248) |
| | **PPI** (µU/ml) | 45.8 (38.3-53.3) | 48.1 (37.9-58.4) | 38.4 (30.4-46.3) |
| | **GSH** (µM) | 416 (353-478) | 468 (392-544) | 515 (438-593) ** |
| | **GSSG** (µM) | 272 (225-320) | 267 (219-315) | 286 (244-327) |
| | **8-OHdG** (ng/µg DNA) | 407.02 (375.77-438.28) | 401.08 (367.41-434.74 ) | 440.62 (408.18-473.06) |
| **DG** | **HbA1c** (%) | 8.6 (8.0-9.2) | 7.5 (7.1-7.9) *** | 7.7 (7.3-8.1) *** |
| | **FPG** (mg/dl) | 150 (135-164) | 134 (123-146) * | 146 (131-161) |
| | **FPI** (µU/ml) | 12.3 (10.3-14.4) | 15.8 (11.3-20.2) * | 14 (10.9-17.1) |
| | **PPG** (mg/dl) | 221 (201-242) | 197 (180-214) * | 212 (192-231) |
| | **PPI** (µU/ml) | 50.8 (35.9-65.7) | 55.1 (42.7-67.6) | 55.4 (43.9-67) |
| | **GSH** (µM) | 452 (366-537) | 1092 (922-1263) *** | 1059 (914-1024) *** |
| | **GSSG** (µM) | 160 (133-187) | 335 (279-391) *** | 291 (236-346) *** |

| | | | | |
|---|---|---|---|---|
| | **8-OHdG** (ng/µg DNA) | 487.81 (465.83-509.79) | 374.8 (341.77-407.83)*** | 278.21 (242.35-314.06) *** |

**Table S5. Biochemical parameters of elderly diabetic subjects at different visits.** Biochemical measurements of elderly diabetic subjects (age > 55 years) in D and DG groups at $\alpha$, β, and $\gamma$visits are shown here with mean and 95% confidence interval. Significance levels (*) displayed at $\beta$, and $\gamma$visits denote the comparisons with $\alpha$ visit using permutation tests. Significance levels are *p < 0.05, **p < 0.01, ***p < 0.001 for respective comparisons (See Table S7 for p-value of comparisons using t-tests and alternate comparisons using t-tests).

**Table S6.**

| Groups | Variables | p-values using permutation tests | | p-values using t-tests | |
|---|---|---|---|---|---|
| | | Between α and β visits | Between α and γ visits | Between α and β visits | Between α and γ visits |
| **Control** | **HbA1c** | 0.966 | $3\times 10^{-4}$ *** | 0.932 | $2.58\times 10^{-4}$ *** |
| | **FPG** | 0.884 | 0.296 | 0.873 | 0.290 |
| | **FPI** | 0.039* | 0.022 * | 0.105 | 0.0235 * |
| | **PPG** | 0.041* | 0.962 | 0.041 * | 0.938 |
| | **PPI** | 0.30 | 0.839 | 0.30 | 0.838 |
| | **GSH** | 0.637 | 0.687 | 0.636 | 0.687 |
| | **GSSG** | 0.0604 | 0.642 | 0.061 | 0.641 |
| | 8-OHdG | 0.488 | 0.88 | 0.487 | 0.882 |
| **D** | **HbA1c** | 0.005 ** | 0.10 | 0.0053 ** | 0.098 |
| | **FPG** | 0.004 ** | 0.242 | 0.0049 ** | 0.238 |
| | **FPI** | 0.032 * | 0.0324 * | 0.037 * | 0.0353 * |
| | **PPG** | 0.068 | 0.248 | 0.068 | 0.246 |
| | **PPI** | 0.251 | 0.695 | 0.248 | 0.693 |
| | **GSH** | 0.255 | $2.94\times 10^{-4}$ ** | 0.255 | $2.96\times 10^{-4}$ *** |

| | | | | | |
|---|---|---|---|---|---|
| | **GSSG** | 0.326 | 0.457 | 0.324 | 0.456 |
| | **8-OHdG** | **0.235** | **0.174** | **0.292** | **0.174** |
| **DG** | **HbA1c** | $1.49 \times 10^{-6}$ *** | $7.12 \times 10^{-4}$ *** | $9.31\times 10^{-6}$ *** | 0.0011 ** |
| | **FPG** | 0.0543 | 0.675 | 0.0549 | 0.671 |
| | **FPI** | 0.994 | 0.989 | 0.983 | 0.757 |
| | **PPG** | 0.193 | 0.587 | 0.192 | 0.582 |
| | **PPI** | 0.910 | 0.299 | 0.910 | 0.288 |
| | **GSH** | $1.1 \times 10^{-19}$ *** | $3.09 \times 10^{-16}$ *** | $4.44\times 10^{-18}$ *** | $4.23\times 10^{-17}$ *** |
| | **GSSG** | $3.33\times 10^{-12}$ *** | $4.86 \times 10^{-8}$ *** | $7.83\times 10^{-11}$ *** | $2.59\times 10^{-7}$ *** |
| | **8-OHdG** | **$2.08\times 10^{-8}$ ***** | **$1.788\times 10^{-14}$ ***** | **$1.6\times 10^{-8}$ ***** | **$1.53\times 10^{-15}$ ***** |

**Table S6. Significance levels of intra-group comparisons between biochemical parameters at different visits.** Biochemical parameters at different visits were compared using permutation tests and t-tests for Control, D, and DG groups. p-values of comparisons are given here in this table. Significance levels are *p < 0.05, **p < 0.01, ***p < 0.001 for respective comparisons.

**Table S7.**

| **Biochemical variables (X)** | **D group** | | **DG group** | |
|---|---|---|---|---|
| | $\Delta\bar{X}^{D}_{\gamma-\alpha}$ **($s_1$)** | $N_D$ | $\Delta\bar{X}^{DG}_{\gamma-\alpha}$ **($s_2$)** | $N_{DG}$ |
| **HbA1c** (%) | -0.1 (1.6) | 44 | -1 (2) | 54 |
| **FPG** (mg/dl) | -12 (59) | 44 | -3(55) | 54 |
| **PPG** (mg/dl) | -1 (105) | 44 | -11(82) | 53 |
| **FPI** (µU/ml) | -2.2 (7) | 44 | 1.6(6.5) | 54 |
| **PPI** (µU/ml) | -3.1(23.7) | 44 | 3.7(42.5) | 53 |
| **GSH** (µM) | 115(230) | 44 | 607 (545) | 53 |
| **GSSG** (µM) | 8(140) | 44 | 1 (7) | 53 |
| **8-OHdG** (ng/µg DNA) | 33.19 (173.35) | 45 | -209.60 (162.64) | 54 |

**Table S7. Six-month biochemical changes in elderly diabetic subjects.** Mean ($\Delta\bar{X}^{D}_{\gamma-\alpha}$ and $\Delta\bar{X}^{DG}_{\gamma-\alpha}$) and standard deviation ($s_1$and $s_2$) of 6-month biochemical changes in elderly subjects of D and DG groups (age > 55 years ) are shown here. $N_D$ and $N_{DG}$ are the number of individuals in D and DG, respectively.

**Table S8.**

| Groups | Variables | p-values using permutation tests | | p-values using t-tests | |
|---|---|---|---|---|---|
| | | Between α and β visits | Between α and γ visits | Between α and β visits | Between α and γ visits |
| **D** | **HbA1c** | 0.788 | 0.622 | 0.776 | 0.605 |
| | **FPG** | 0.028 * | 0.198 | 0.0293 * | 0.194 |
| | **FPI** | 0.032 * | 0.042 * | 0.0480 * | 0.0492 * |
| | **PPG** | 0.758 | 0.963 | 0.753 | 0.961 |
| | **PPI** | 0.587 | 0.397 | 0.582 | 0.394 |
| | **GSH** | 0.143 | 0.002 ** | 0.143 | 0.0019 ** |
| | **GSSG** | 0.811 | 0.698 | 0.808 | 0.697 |
| | **8-OHdG** | 0.784 | 0.205 | 0.784 | 0.206 |
| **DG** | **HbA1c** | $5.25 \times 10^{-6}$ *** | $7.2 \times 10^{-4}$ *** | $3.78\times 10^{-5}$ *** | 0.0018 ** |
| | **FPG** | 0.0136 * | 0.669 | 0.0148 * | 0.665 |
| | **FPI** | 0.046 * | 0.069 | 0.0757 | 0.0689 |
| | **PPG** | 0.013 * | 0.324 | 0.0136 * | 0.321 |
| | **PPI** | 0.523 | 0.528 | 0.521 | 0.524 |
| | **GSH** | $6.51 \times 10^{-10}$ *** | $2.31 \times 10^{-10}$ *** | $2.77\times 10^{-9}$ *** | $8.27\times 10^{-11}$ *** |
| | **GSSG** | $2.94 \times 10^{-8}$ *** | $8.71 \times 10^{-6}$ *** | $3.75\times 10^{-7}$ *** | $4.28\times 10^{-5}$ *** |
| | **8-OHdG** | 3.62 × | $4.43\times 10^{-11}$*** | $2.22\times 10^{-7}$*** | $5.44\times 10^{-13}$*** |

| | | |
|---|---|---|
| | | $10^{-7}$*** |

**Table S8. Significance levels of intra-group comparisons between biochemical parameters at different visits of elderly subjects.** Biochemical concentrations at different visits of elderly subjects (age>55 years) in Control, D, and DG groups were compared using permutation tests and t-tests. p-values of comparisons are shown in this table. Significance levels are *p < 0.05, **p < 0.01, ***p < 0.001 for respective comparisons.
